# Machine learning enabled prediction of digital biomarkers from whole slide histopathology images

**DOI:** 10.1101/2024.01.06.24300926

**Authors:** Zachary R McCaw, Anna Shcherbina, Yajas Shah, Davey Huang, Serra Elliott, Peter M Szabo, Benjamin Dulken, Sacha Holland, Philip Tagari, David Light, Daphne Koller, Christopher Probert

**Affiliations:** Insitro, South San Francisco, CA, USA; Department of Pathology, Stanford School of Medicine, Stanford, CA, USA

**Author notes:** Corresponding authors: DK, CP. Equal contribution (Zachary R McCaw, Anna Shcherbina).

## Abstract

Current predictive biomarkers generally leverage technologies such as immunohistochemistry or genetic analysis, which may require specialized equipment, be time-intensive to deploy, or incur human error. In this paper, we present an alternative approach for the development and deployment of a class of predictive biomarkers, leveraging deep learning on digital images of hematoxylin and eosin (H&E)-stained biopsy samples to simultaneously predict a range of molecular factors that are relevant to treatment selection and response. Our framework begins with the training of a pan-solid tumor H&E foundation model, which can generate a universal featurization of H&E-stained tissue images. This featurization becomes the input to machine learning models that perform multi-target, pan-cancer imputation. For a set of 352 drug targets, we show the ability to predict with high accuracy: copy number amplifications, target RNA expression, and an RNA-derived “amplification signature” that captures the transcriptional consequences of an amplification event. We facilitate exploratory analyses by making broad predictions initially. Having identified the subset of biomarkers relevant to a patient population of interest, we develop specialized machine learning models, built on the same foundational featurization, which achieve even higher performance for key biomarkers in tumor types of interest. Moreover, our models are robust, generalizing with minimal loss of performance across different patient populations. By generating imputations from tile-level featurizations, we enable spatial overlays of molecular annotations on top of whole-slide images. These annotation maps provide a clear means of interpreting the histological correlates of our model’s predictions, and align with features identified by expert pathologist review. Overall, our work demonstrates a flexible and scalable framework for imputing molecular measurements from H&E, providing a generalizable approach to the development and deployment of predictive biomarkers for targeted therapeutics in cancer.

## 1 Introduction

Cancer is a highly heterogeneous disease, and despite significant advances in the discovery and development of “precision” approaches to management, patients’ responses to targeted treatments can still be highly variable, without an understanding as to why (see for example [1, 2]). Growth in the development of targeted therapies has accelerated the use of predictive biomarkers to identify the patients that are more likely to respond to a drug. Indeed, studies have shown that oncology trials that use biomarkers have a considerably higher success rate [3, 4], with a nearly 5-fold increased likelihood of drug approval across all indications combined, and a 12-, 8- and 7-fold improvement for breast cancer, melanoma and non-small cell lung cancer (NSCLC), respectively.

Current predictive biomarkers generally leverage one of several assay types: immunohistochemistry (IHC) on biopsy slides; genetic analysis, including karyotyping, fluorescence in situ hybridization, and DNA sequencing; or transcript levels of a small set of genes, measured either via polymerase chain reaction (PCR) or (rarely) broad-based RNA sequencing. The development and deployment of these approaches present significant challenges. First and foremost, these methods rely on specialized assays that are not universally available across cancer centers, and even more so in resource-poor settings. Second, some of these technologies such as IHC, require manual assessment by a trained individual, which might increase variability and decrease reproducibility of the assay results. Third, they entail additional cost and, even more importantly, require additional time that could postpone the time to a diagnosis and the initiation of treatment. Moreover, while the sequencing-based assays leverage technologies that have seen generally broad adoption and their use is fairly standardized, a biomarker that utilizes targeted staining or probes, including IHC and PCR, will usually require the development and extensive testing of specialized assays and reagents.

Current development paradigms and available technologies favor early selection of a biomarker, at a stage where available data is generally based on poorly representative preclinical models and/or underpowered phase 1 studies, both of which fail to capture the heterogeneity in human patient populations. This drives a tendency towards simple biomarkers that are largely driven by human mechanistic understanding of the disease, usually either genetic aberrations as measured by sequencing, or transcript/protein expression as measured by chemistry. As a consequence, the labeled population is often overly restricted – reducing the set of patients who benefit – or overly broad – subjecting a subset of patients to a drug that has limited efficacy, while still carrying the risk of toxicity and delaying treatment with a potentially more efficacious therapy.

In this paper, we present an alternative approach for the development and deployment of a class of predictive biomarkers, leveraging deep learning on images of hematoxylin and eosin (H&E)-stained biopsy samples to simultaneously predict a range of molecular factors that are relevant to treatment selection and response. Such images are routinely collected and processed for almost all solid tumor patients (world-wide), and are increasingly digitized [5, 6, 7, 8]. As has been recently shown, these images are incredibly rich in information, and enable automated disease detection, prognosis prediction, cancer grading, histological and molecular subtyping, and customized treatment planning [9]. In particular, multiple papers have demonstrated the ability to predict molecular biomarkers from H&E images, mostly focusing on therapeutically-relevant, recurrent somatic alterations [10, 11, 12, 13, 14, 15], but more recently extending to other molecular modalities, including RNA expression, protein abundance, and more [16].

Existing studies have predominantly utilized a per-task supervised learning framework, where a single deep learning model is trained to predict a specific, defined biomarker in clinical use, in a given type of cancer, directly from the H&E images. This approach limits the usable training data to individuals within a single cancer for whom the known biomarker has been measured. In this work, we develop a novel multi-cancer, multi-biomarker prediction framework that enables us to leverage the commonality of cancer mechanisms that manifest both in the H&E slides and in the molecular readouts. Specifically, we begin by training a pan-solid tumor H&E foundation model (similar to [9, 17, 18]; also see [19]), learning a universal featurization of tissue H&E images.

We then use the foundational embeddings as input to downstream ML models. By predicting multiple biomarkers simultaneously using a multi-task learning approach, our first set of downstream models allow for exploratory analysis and broad discovery. To enhance interpretability, these models predict on the basis of tile- rather than slide-level featurizations, where a tile constitutes a small element of the far larger whole slide image. Having tile-level predictions enables the generation and overlay of annotation maps, highlighting the regions of a slide driving the model’s predictions. We show that despite being trained on bulk, rather than spatially resolved, molecular data, our model is able to learn spatial variation in tumor cell molecular markers that correlates with the regions identified as cancerous by blinded pathologist review.

Our initially broad set of imputations allows for hypothesis-free investigations of which biomarkers are relevant to which patient populations, and for the identification of biomarkers that differentiate patient subgroups. Once a smaller set of biomarkers specific to the patient population of interest has been selected from the discovery panel, we show that specialized models can be trained, starting from the same foundational featurization, which can surpass the discovery model at predicting key biomarkers in targeted subgroups. This two step process of imputing broadly then specializing on a more-focused subset enables both the discovery of novel biomarkers and the optimization of their diagnostic performance.

To demonstrate the value of this method, we focus on biomarkers that are relevant to the efficacy of drugs whose mechanism of action (MOA) is based on the differential recognition and killing of cancer cells via the abundance of a particular protein target: antibodies (both mono-specific and multi-specific), antibody-drug conjugates (ADCs), and T-cell engagers. For relevant targets, we explore three potential imputed biomarkers: copy-number amplification (CNAs), RNA transcript level, and an RNA-derived signature capturing the effect of a target’s CNA on the transcriptome. Across a large and diverse set of cancer types and biomarkers, our methods deliver high-accuracy patient-specific predictions of molecular readouts, both for continuous and dichotomized versions of these biomarkers. We also show that our ability to provide accurate predictions is considerably greater for RNA transcript levels than for CNAs, which further supports our use of RNA-derived signatures as proxies for CNAs. As such, our work offers the opportunity to optimize the patient population for a targeted therapy beyond the use of genetic alterations or IHC, but without going to the other extreme of an overly broad label covering an excessively heterogeneous patient set. Moreover, despite the fact that our imputation models were trained on bulk readouts, they enable us to overlay spatially varying, tile-level predictions on top of the input histology images, providing a lens of interpretability and enabling clinicians to gauge (e.g.) if pairs or sets of biomarkers are spatially colocalizing within a tumor. Overall, our results support the viability and ongoing exploration of using highly scalable molecular predictions from H&E as a flexible and generalizable approach to the development and deployment of predictive biomarkers for targeted therapeutics in cancer.

## 2 Methods

### 2.1 Data Sets and Data Preparation

#### Data Sets

Our study employed data from The Cancer Genome Atlas (TCGA), a public research resource that includes genetic, molecular, and histological data from 11K patient and over 20K primary tumors across 33 cancer types [20]. Molecular and histological data from an additional 2.6K patients were obtained from a commercially available multi-center cancer research resource (cohort A).

#### Target Gene Identification

A commercial pharmaceutical database was queried to identify drugs whose therapeutic class was labeled as antibody-drug conjugate (ADC), T-cell engager, or antibody, including both mono-specific and multi-specific antibodies. For ADCs and T-cell engagers, drugs at any stage of development were retained, while for antibodies (a larger class), drugs whose development had ceased were excluded. The overall list of drugs was filtered to those with specified targets. Each remaining drug was mapped to a HGNC gene symbol, and the union of all gene symbols was taken, resulting in 352 unique targets.

#### Copy Number Amplification Calling

Two approaches were utilized to identify genes with focal amplifications based on whole exomes from tumor specimens: GISTIC2 [21] (v2.0.22), which estimates copy number relative to a matched normal sample, and Sequenza [22] (v3), which estimates the absolute copy number. A gene was designated as focally amplified if it received a GISTIC2 score of 2, or if it exhibited a copy number greater than twice the ploidy based on Sequenza.

#### Expression Data Preprocessing

Augmented TCGA STAR+RSEM gene counts for 11,155 samples, generated from the Genomic Data Commons (GDC) [23] standard pipeline and aligned to GRCh38, were downloaded from the GDC portal on 07/13/2023. A corresponding STAR+RSEM gene count matrix for 2,733 samples was prepared in cohort A. Transcript per million (TPM) matrices were concatenated, filtered to genes with non-trivial expression (requiring TPM *>* 1 in at least one subject), and subset to protein-coding genes from Gencode V43 [22], resulting in 19,421 unique genes. The resulting TPM matrix was log2-transformed then quantile normalized via the limma voom function [24] in R [25]. Subsequently, edgeR’s removeBatchEffects function [26] was applied to regress out the cohort effect (TCGA vs. cohort A). The resulting log_2_(TPM) matrix was assessed for possible batch effects from sequencing instrument and sequencing center via principal component analysis and lmfit on potential batch drivers. No other significant batch effects were identified. This joint expression matrix was used as input for downstream analyses.

#### Histology Preprocessing

A total of 30,032 hematoxylin and eosin (H&E)-stained whole slide images (WSIs), corresponding to 11,428 unique patients, were downloaded from the GDC. The tissue-bearing foreground of the image is extracted, and low-frequency super-cellular artifacts such as tissue folds, out-of-focus regions, and pen markings removed, using WSI Spectral Thresholding for Artifact Removal (WSI-STAR), an internally-developed histology slide quality control procedure currently under review. To account for differences in staining protocols across studies centers, color channels were normalized using Macenko’s method [27]. Each slide was divided into non-overlapping 256 *×* 256 tiles at a resolution of 1 *µ*m per pixel (MPP), and tiles were filtered to those with at least 90% foreground, resulting in 180M individual tiles. WSIs for 1,000 patients from cohort A were processed in the same manner, resulting in 8M tiles.

### 2.2 Amplification Signatures

#### Differential Expression Analysis

Differential expression analysis between copy-number amplified and copy-number normal (i.e. diploid) patients was performed using the limma voom package in R. For each amplification, limma models (*∼* CNA status + cohort) were fit to identify deferentially expressed genes with a false discovery rate (FDR)-corrected p-value (i.e. q-value) *<* 0.01 and an absolute log_2_ fold change *>* 0.3. Such models were fit jointly to the TCGA and cohort A data sets. No significant differences were observed in a sensitivity analysis where the TCGA and cohort A data sets were analyzed separately.

#### Amplification Signature Construction

An expression signature for each amplification was constructed by taking the dot product of the observed expression levels of differentially expressed genes with a set of weights. In detail, for amplification *k*, suppose there were *J_k_*differentially expressed genes. Let *G_ij_* denote the observed expression level of gene *j* in subject *i*. The signature for subject *i* with respect to the *k*th amplification was:

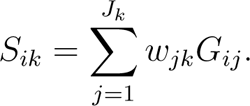

The weight *w_jk_*was the sign of the log_2_ fold change of gene *j* in amplification *k* scaled by the absolute log_10_ q-value. This scheme affords more weight to genes where there is greater evidence of differential expression. A more positive signature *S_ik_*indicates that subject *i* has an expression profile consistent with amplification *k*, even if that subject did not have amplification of *k* based on copy number analysis. Below, we develop models to impute expression levels and amplification signatures. While in principle the amplification signature could be calculated from imputed expression levels, in practice better performance is obtained by developing models to directly impute the amplification signature (using labels derived from observed expression levels).

### 2.3 Machine Learning Analysis

#### Histology Embedding Generation

We trained a vision transformer [28] (ViT)-type model on 3M randomly selected 256 *×* 256 1MPP tiles from TCGA using the self-supervised <u>di</u>stillation with <u>no</u> labels (DINO) [29] algorithm. Briefly, given a collection of unlabeled images, DINO trains a student network (i.e. the ViT) to match the output of a teacher network. This task is made more challenging by the fact that the student and teacher networks receive different “views” of the input image. Training was monitored by periodically evaluating the utility of embeddings extracted from the teacher network for several downstream tasks, including cancer subtype classification and overall survival prediction, within an independent validation set of 100K tiles. Tile-level embeddings generated by the final model served as the inputs to downstream modeling tasks.

#### Biomarker Foundation Model Training

Three sets of neural network models were trained via 8-fold cross validation to predict gene expression, copy number amplification, and gene signatures from H&E tile embeddings of dimension 768. The training was performed in pytorch (2.1.0). Two major architecture classes were used. The first, a 4-layer sequential network consisting of linear layers interspersed with ReLU and dropout layers, is reproduced below. Training and evaluation data was fed to the model in batches of size 2000.

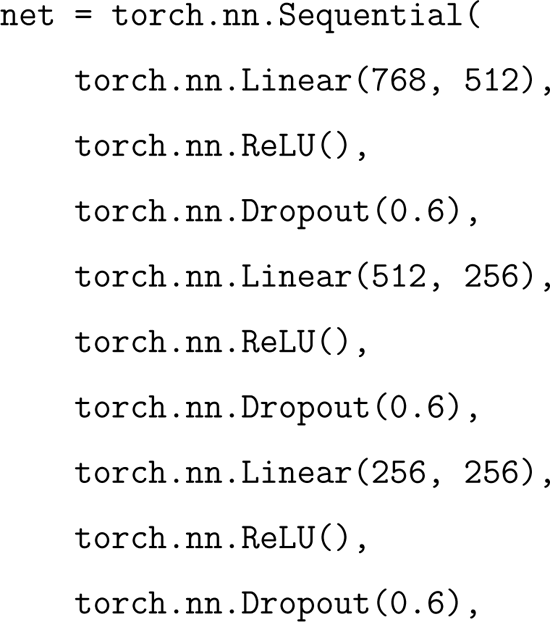

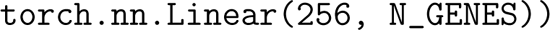

The second class of model extended the first to include cross-tile attention, building on the transMIL [30] architecture, using batch size of 1 with gradient accumulation for 400 batches. Optimization was performed using Adam [31], starting with a learning rate of 1e-4, which decayed exponentially (`gamma`=0.96) after 2 consecutive epochs of no improvement. An early stopping threshold of 3 or 4 (depending on the model) consecutive epochs with no improvement in validation loss was utilized to indicate training completion.

For regression tasks (predicting target expression or the amplification signature), the objective was Huber loss with `delta`=1.0. For classification tasks (predicting copy number amplification, elevated target expression, or elevated amplification signature), the objective was binary cross entropy loss, with the minority (positive) class inversely weighted by class prevalence. When performing classification for elevated target expression or amplification signature, the positive class was defined as those patients exceeding the 95% percentile (p95). Label smoothing [32] was applied during training, with p0-p50 assigned a label of 0; p50-p90 a label of 0.1; p90-p95 a label of 0.9; and p95-p100 a label of 1.0. Label smoothing was not possible in the case of CNA labels, which are intrinsically binary.

#### Biomarker Specialized Model Training

Building on the promising results from the expression and signature classification foundation models, specialized models were trained to predict MET expression and signature within the NSLC and COAD cohorts. Training was performed on tile-level H&E embeddings using the architecture:

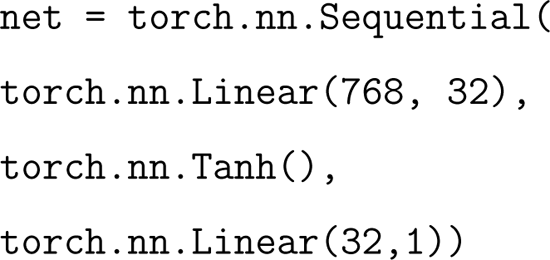

Inputs to the model were restricted to H&E data from the cohort of interest (NSLC or COAD), keeping the same subject splits as in the foundation models to avoid contamination, but pruning out training and evaluation subjects from other cohorts.

Training was performed via a binary cross entropy loss using the Adam optimizer with a weight decay of 1e-4, a learning rate 0.001, and early stopping enabled after three consecutive epochs of no reduction in evaluation loss. Binarization and label smoothing was performed as described above for the foundation models.

#### Assessing Model Performance

Model performance was evaluated via an 8-fold cross-validation procedure, where the model was trained on 7 folds and the evaluated on the held-out fold. For regression tasks, the evaluation metrics included the Pearson and Spearman correlations. For classification tasks, the evaluation metrics included the area under the precision-recall curve (AUPRC) and the area under the receiver operating characteristic (AUROC). While the models emit tile-level predictions, tiles are clustered within patients, and the labels are patient level. Performance metrics were aggregated from tile to patient level by taking the mean. For pan-cancer analyses, performance is assessed across all patients, whereas for stratified analyses, performance is assessed first within each cancer type, than averaged across cancer types. The stratified analysis restricts to cancer types with at least 100 available patients to ensure the performance metrics can be estimated with reasonable precision. Due to the low prevalence (e.g. *<* 1%) of certain CNAs, for stratified CNA analyses, on targets where at least 3 patients harbored CNAs in a given cancer type are included.

### 2.4 Survival Analysis

Curated OS labels for patients in TCGA were obtained from [33]. Within cohort A, therapy-specific OS was defined as the time from therapy onset to the patient’s death. In instances where no death report was available, patients were censored at the time of last follow-up. Analyses were performed in cohort A, where more-detailed clinical data were available. Hazard ratios quantifying the association between OS and predicted biomarkers were estimated via the Cox proportional hazards model [34], adjusting for the age at diagnosis, age at disease staging, pre-treatment stage, sex, cancer type, metastatic status, number of unique prior therapies, and time from diagnosis to treatment with the therapy of interest. Patients were partitioned into two groups (“high” and “low”) on the basis of their amplification signature, but without reference to their survival. The significance of differential survival between these groups was assessed via the HR from the Cox model. Adjusted Kaplan-Meier curves were calculated using the direct standardization approach [35, 36].

## 3 Results

### 3.1 Biomarker Prediction

#### CNA Prediction

Copy number amplifications (CNAs) were called for each of 352 target genes (hereafter, “targets”). Within the overall cohort (*n* = 14, 007), the median amplification prevalence was 1.1% (range: 0.2% to 7.8%; also see **Supplemental Figure 1**). We developed multi-task, binary-outcome models to simultaneously predict CNA status for all 352 targets. The inputs to these prediction models were embeddings of 256 by 256, 1 *µ*m per pixel tiles from digital, whole-slide histopathology images (WSIs). Patient-level predictions are obtained by taking the mean across all tiles within a patient’s WSIs. Here and throughout, patient predictions are generated using an 8-fold cross-validation (CV) procedure such that the model which generates a patient’s prediction does not see that patient’s data during training.

Figure 1 presents the distribution of AUROCs, across targets, and mean AUROC in each of 26 cancer types with at least 100 patients. Due to the low prevalence of certain CNAs, when assessing performance within cancer types, metrics are only reported for those targets where at least 3 patients harbored amplifications. The mean AUROC across targets is summarized in **Table 1**. The heatmaps in **Supplemental** Figure 2 present, for each target, the area under the receiver operating characteristic (AUROC) for predicting CNAs, stratified by cancer type. The pan-cancer analysis evaluates performance across all available patients.

**Figure 1:**
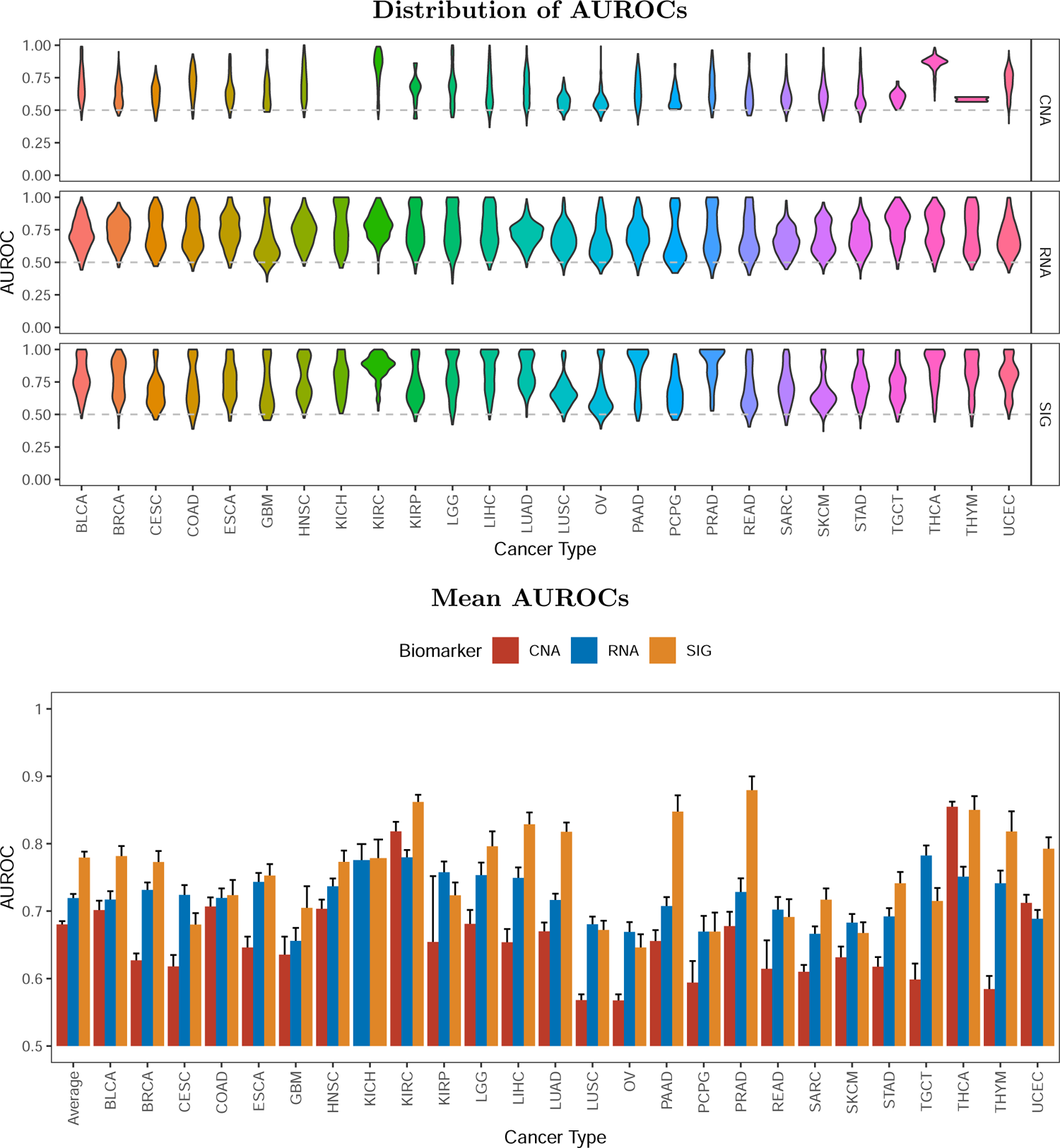
Cross-modality comparison of binary digital biomarker prediction quality, stratified by cancer-type. Performance is evaluated at the patient level in a held-out evaluation set and averaged across 8 cross-validation folds. Metrics are calculated separately in each cancer type with *≥* 100 patients. Within cancer types, CNA results are only presented for targets where at least 3 patients harbored an amplification. The mean and distribution are shown across up to 352 target genes. For CNA, the task was to predict whether the patient harbored an amplification. For target expression (RNA) and the amplification signature (SIG), the task was to predict whether the patient’s expression/signature level exceeded the 95th percentile. Error bars are 95% confidence intervals.

**Table 1:** Overall performance for biomarker prediction from digital histopathology. Performance is evaluated at the patient level in a held-out evaluation set and averaged across 8 cross-validation folds. The 3 types of biomarkers are copy number amplification (CNA), target expression level (RNA), and amplification signature score (SIG). For binary classification, the area under the receiver operating characteristics (AUROC) is shown. For regression, Spearman correlation between the observed and predicted values is shown.

| Cohort | N | Biomarker | AUROC | Spearman |
| --- | --- | --- | --- | --- |
| Pan-cacner | 14,007 | CNA | 0.734 |  |
| Pan-cacner | 14,007 | RNA | 0.853 | 0.628 |
| Pan-cacner | 14,007 | SIG | 0.897 | 0.665 |
| Stratified | 12,328 | CNA | 0.680 |  |
| Stratified | 12,328 | RNA | 0.719 | 0.318 |
| Stratified | 12,328 | SIG | 0.779 | 0.333 |
| Breast | 1,455 | CNA | 0.627 |  |
| Breast | 1,455 | RNA | 0.731 | 0.372 |
| Breast | 1,455 | SIG | 0.773 | 0.418 |
| Colorectal | 629 | CNA | 0.707 |  |
| Colorectal | 629 | RNA | 0.720 | 0.324 |
| Colorectal | 629 | SIG | 0.724 | 0.293 |

The stratified analysis evaluates performance separately in each cancer type, then takes the mean across cancer types. The distinction between these approaches is that the stratified analysis evaluates how well the model learns to differentiate CNA risk within cancer types, whereas the pan-cancer analysis examines how well the model learns to differentiate risk both within and between cancer types. To demonstrate the within-cancer-type performance, the performance within two specific cohorts of interest, breast and colorectal, is also presented.

#### Expression Prediction

Previous work has associated copy number amplification with differential gene expression across cancer types [37]. **Supplemental** Figure 4 presents the differences in expression between patients with and without CNAs across 347 targets with available RNA. Of these, 207 (59.7%) were significantly differentially expressed, the vast majority (197/207; 95.2%) having higher mean expression in patients with amplifications. We hypothesized that, by providing a continuous supervisory signal, modeling RNA would enable us to train more accurate biomarker prediction models.

We therefore developed multi-task, continuous-outcome models to simultaneously predict, on the basis of histopathology tile embeddings, the expression levels for 347 of the 352 targets with available RNA. Figure 2 compares the observed and predicted expression matrices pan-cancer. Analogous matrices subset to breast and colorectal cancer are presented in **Supplemental** Figure 5. Figure 3 presents the distribution of correlations, across targets, between a patient’s observed and predicted expression levels, and the mean correlation by cancer type. Specifically, patient level predictions were first generated via 8-fold CV, then for each target, the correlation between the observed and predicted expression levels was calculated across patients. Pan-cancer, the mean cross-validated Spearman correlation was 62.8%, and stratifying by cancer type, the mean Spearman correlation was 31.8% (**Table 1**). As expected, correlations are higher in the pan-cancer analysis, where the model benefits from learning to distinguish differences both within and between cancer types. **Supplemental** Figure 9 presents the correlations for all targets broken down by cancer type.

**Figure 2:**
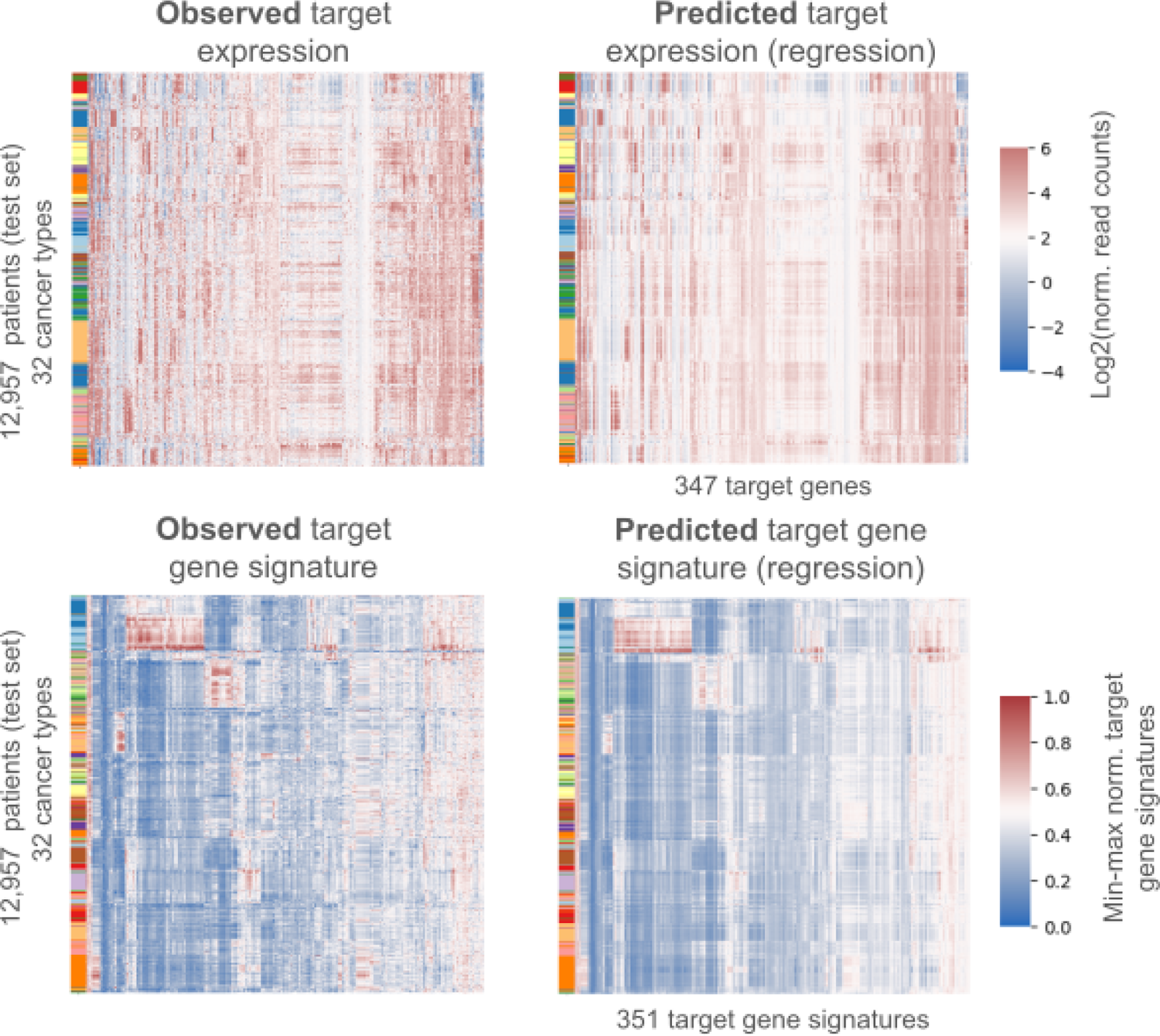
Comparison of observed expression and signature matrices with those predicted on the basis of histopathology. Predictions are generated via cross-validation, such that a patient is not used to train the model that generates their predictions. Left is the observed gene expression or signature matrix. Right are prediction based on digital pathology. The color within the heatmap describes the level of expression or magnitude of the amplification signature. The color bar on the left of each plot annotates cancer type.

**Figure 3:**
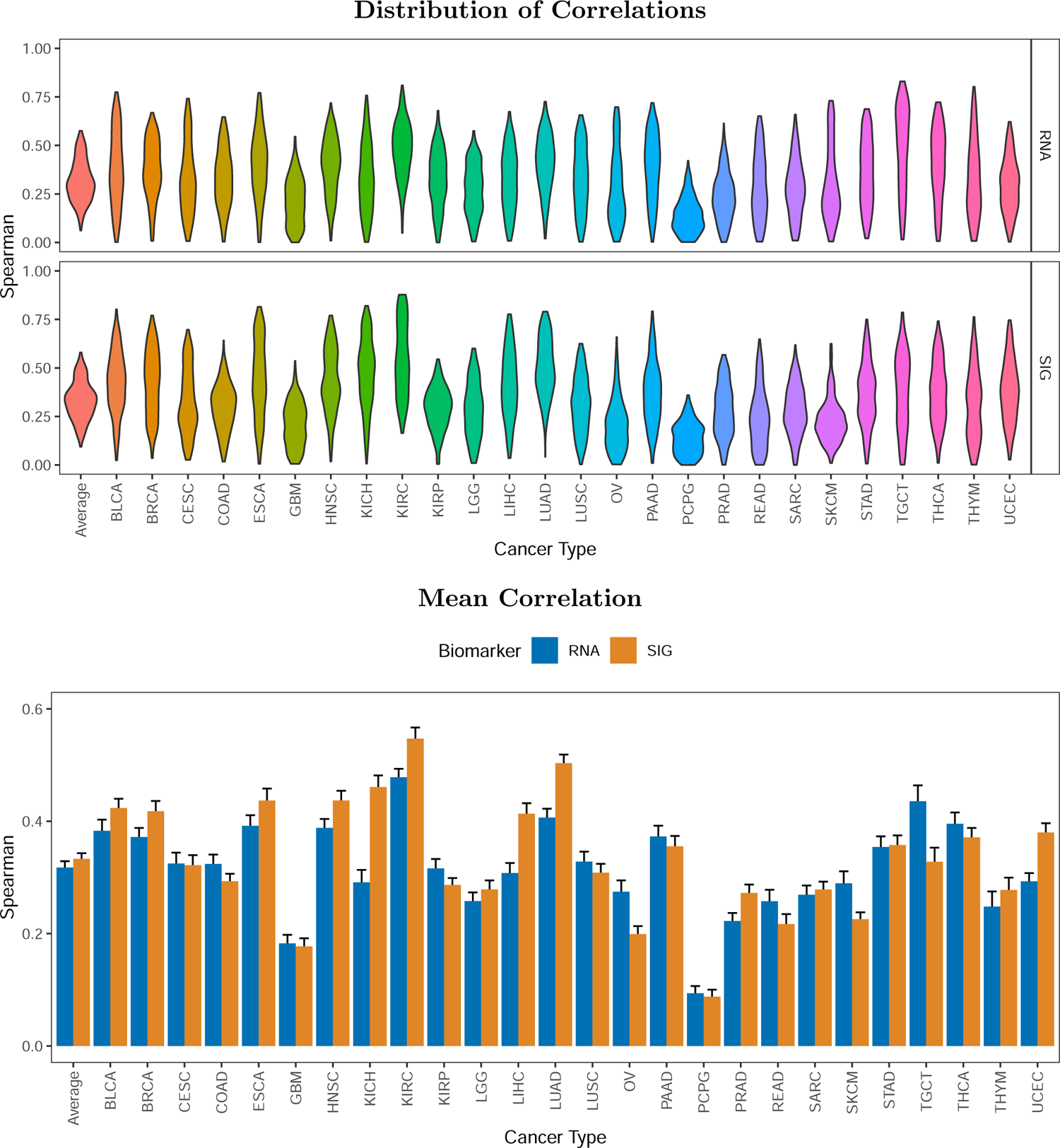
Cross-modality comparison of continuous digital biomarker prediction quality, stratified by cancer-type. Performance is evaluated at the patient level in a held-out evaluation set and averaged across 8 cross-validation folds. Metrics are calculated separately in each cancer type with *≥* 100 patients. The distributions are shown across up to 352 target genes. For RNA, the task is to predict the normalized log_2_ expression level. For SIG, the task is to predict the min-max normalized amplification signature.

To enable comparison with the binary CNA prediction task, we dichotomized the expression of each target at its 95th percentile (p95), and developed multi-task binary-outcome models to predict whether a patient’s expression exceeded the p95, suggesting that the target was highly expressed. **Supplemental** Figure 6 presents results for all targets stratifying by cancer type, and Figure 1 presents the distributions and means for each cancer type. Consistent with our hypothesis, elevated target expression was generally more predictable than CNA status. As shown in **Table 1** the pan-cancer AUROC increased from 73.4% to 85.3%, and the stratified AUROC increased from 70.0% to 71.9%.

#### Amplification Signatures

As discussed above (see **Supplemental** Figure 4), there is only modest concordance between CNA and differential expression. We reasoned that a broader transcriptional signature capturing changes in expression beyond those of the target gene alone might provide a better predictor of target CNA. For each of the 352 targets, we identified all genes differentially expressed between patients with and without amplifications, and utilized the differentially expressed genes to construct an RNA-based *amplification signature* (for 1 gene, no differentially expressed genes were identified). The amplification signature is a linear combination of expression levels weighted by the magnitude of evidence for differential expression (see Section 2.2). Signatures were min-max normalized to the unit interval for ease of comparison. **Supplemental** Figure 18 presents the distribution of signature scores in patients with and without amplifications. Relative to those without amplifications, the mean signature scores of patients with amplifications were 46.3% higher (**Supplemental Table 3**). For all 351 amplification signatures, there was at least nominally significant evidence, via the Wilcoxon rank-sum test, of differential scores between patients with and without CNAs (median P-value: 1.4 *×* 10*^−^*^27^). **Supplemental** Figure 19 depicts the distribution of correlations, across targets and pan-cancer, between the amplification signature and expression of the amplified gene. In general, the correlation was low, with median *R*^2^ of only 2.0% (**Supplemental Table 4**).

Building on our work predicting target expression, we developed multi-task, continuous-outcome models, analogous to our expression models, for predicting the 351 amplification signatures from histopathology tile embeddings. Figure 3 presents the distribution of correlations, across targets, between a patient’s observed and predicted amplification signatures, and the mean correlation by cancer type. As shown in **Table 1**, predictions of the amplification signature were, on average, more accurate than predictions of target expression. Pan-cancer, the mean cross-validated Spearman correlation was 66.5%, and stratifying by cancer type, the mean Spearman correlation was 33.3% (**Table 1**). **Supplemental** Figure 9 presents the correlations for all targets broken down by cancer type.

We likewise created a binary prediction task, wherein the goal was to predict whether a patient harbored an elevated amplification signature, by dichotomizing each amplification signature at its p95. The distribution and mean AUROC across target, stratified by cancer type, are shown in Figure 1. Pan-cancer, the mean AUROC was 89.7%, and stratified by cancer type, the mean AUROC was 77.9% (**Table 1**). Figure 4 presents the number of targets predicted with an AUROC exceeding a given threshold for the CNA, target expression, and amplification signature binary classification tasks, and **Supplemental Table 2** summarizes the counts exceeding various cutoffs. For example, CNAs in 142 targets, elevated expression in 339 targets, and an elevated signature in 335 targets can be predicted with AUROC exceeding 0.75, pan-cancer. The corresponding counts for the stratified analysis are 25, 97, and 201 for CNA, expression, and signature, respectively. Note that achieving an AUROC of 0.75 or greater in the stratified analysis is a considerably higher bar, as this requires the model to differentiate risk within each cancer type, and do so effectively across many cancer types.

**Figure 4:**
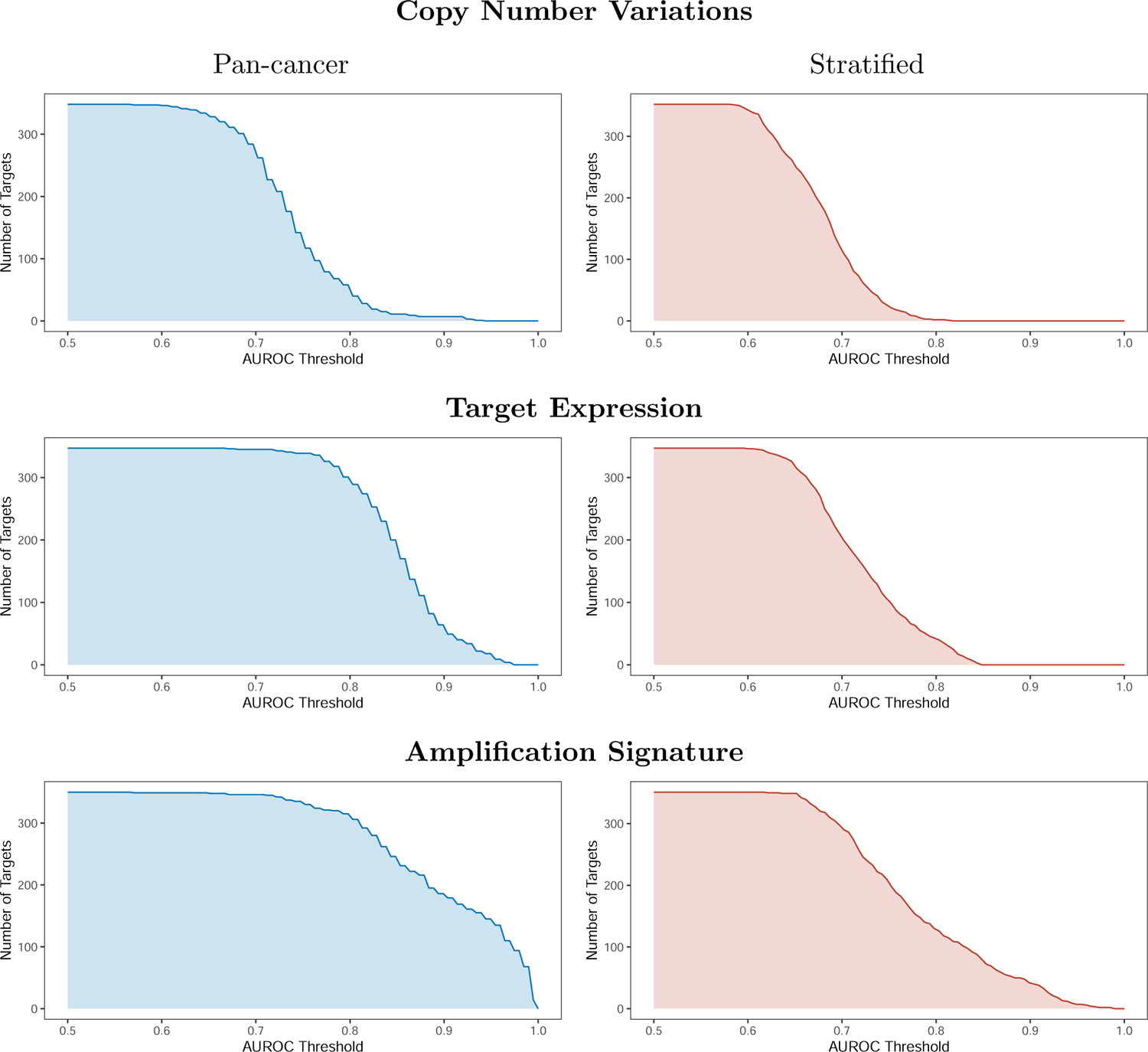
Count of targets with AUROC exceeding a given threshold for the pan-cancer and stratified binary classification task. Performance is evaluated at the patient level in a held-out evaluation set and averaged across 8 cross-validation folds. For the pan-cancer evaluation (left), the AUROC is calculated across all patients. For the stratified evaluation (right), the AUROC is calculated separately within each cancer type, then averaged across cancer types. For copy number amplification, the task was to predict whether the patient harbored an amplification. For target expression and the amplification signature, the task was to predict whether the patient’s expression/signature level exceeded the 95th percentile.

### 3.2 Use Cases

We next sought to assess the utility of our model in a number of distinct applications. Since our study design was based on a set of therapeutically relevant targets, we took a target-driven perspective in exploring use cases.

### *MET* Case Study

We began by looking at *MET*, a target for which there are multiple therapies available and under development. Copy number alteration of *MET* has been associated with worse overall survival across tumor types [38], and specifically in non-small-cell lung cancer [39, 40]. The standard assessment of whether a patient is eligible for a *MET* -targeting therapy utilizes an IHC-based biomarker; indeed, the ADC telisotuzumab vedotin has obtained FDA Breakthrough Therapy Designation for patients with high levels of *MET* overexpression [41]. However, *MET* IHC has been shown to have poor concordance with *MET* CNA [42]. Additionally, it is well established that mechanisms other than CNA that drive *MET* over-expression often also give rise to worse outcomes [43], and are generally much more common than amplification events. For example, in NSCLC, *MET* overexpression is found in 25% – 75% of cases, whereas amplification occurs only in about 4% [43]. Therefore, there is ample opportunity for the development of better biomarkers for identifying patients eligible for *MET* -targeting therapies.

The prevalence of *MET* amplifications in our overall cohort is 1.8%. We first investigated the performance of our core models, as described above, predicting *MET* overexpression and an elevated *MET* amplification signature. As shown in Figure 5(a), pan-cancer, we achieved an AUROC of 0.91 for predicting MET overexpression, and of 0.84 for predicting an elevated amplification signature. Within NSCLC, the pan-cancer model achieved AUROCs of only 0.69 and 0.78 for predicting overexpression and an elevated amplification signature respectively.

**Figure 5:**
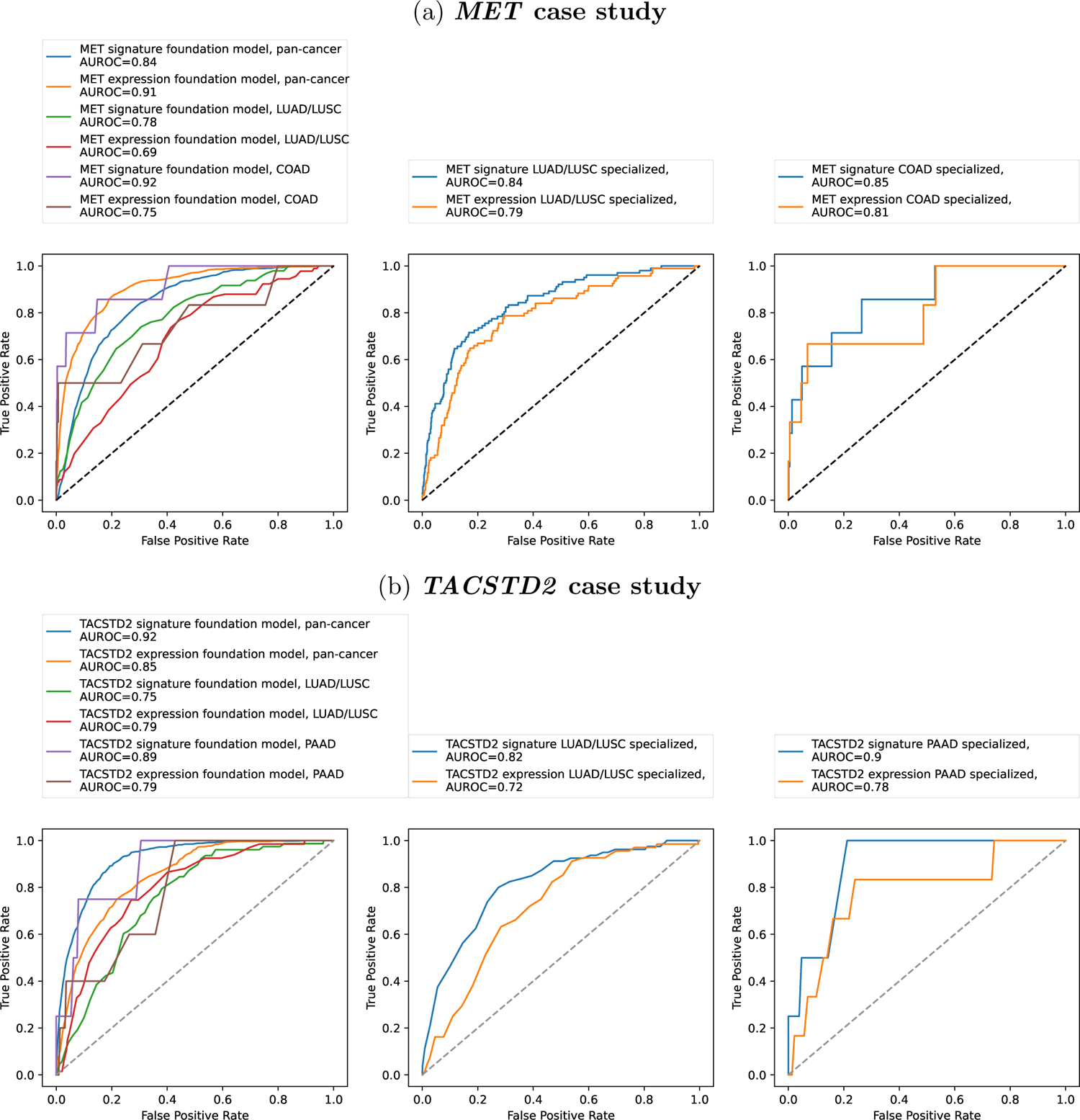
Tailored prediction of overexpression and amplification for *MET* and *TACSTD2*. Left shows performance of models trained to predict overexpression or an elevated amplification signature from the foundation model’s embeddings. Center shows the performance when the model is further specialized for prediction within NSCLC. Right shows performance of a model trained for colorectal cancer prediction, in the case of *MET*, and pancreatic cancer prediction, in the case of *TACSTD2*.

We reasoned that we could further improve the performance within these specific cohorts by specializing our models, that is, training the predictive component of the model on these specific cohorts. Indeed, when models were trained within NSCLC patients specifically, we obtained an AUROC of 0.58 for predicting *MET* amplification, 0.79 for overexpression, and 0.84 for predicting an elevated amplification signature. Our model was also able to predict quantitative MET expression with correlation of 0.38. In work done contemporaneously with ours, Ingale *et al* [44] similarly predict *MET* overexpression in NSCLC; they were able to train on a much larger cohort of NSCLC patients – 605 *MET* + patients versus 38 for us – but used the typical supervised training approach with a single-task model. Their method achieved an AUROC of 0.74 (compared to our 0.79 for overexpression prediction above), but in an artificially balanced test cohort, with equal numbers of cases and controls; in this regime, our approach provided an AUROC of 0.87. We also tested whether our biomarkers have the potential to increase the set of patients where we predict increased MET activity. Indeed, whereas MET CNA identified only 38 NSCLC patients, MET RNA overexpression identified 72 patients, and the MET amplification signature identified 88 patients.

Our pan-cancer approach can also be used to identify new opportunities for biomarker deployment. In particular, our analysis revealed a strong performance in predicting *MET* in colorectal cancer. Although *MET* amplification is rare in colorectal cancer [45, 46], previous work has noted that *MET* overexpression is more common [47, 48], and is prognostic of poorer survival outcomes [49]. We therefore similarly trained a specialized model for colorectal cancer; as shown in Figure 5, this model achieves an AUROC of 0.81 for predicting *MET* overexpression, and of 0.85 for predicting an elevated amplification signature.

### *TACSTD2* Case Study

Antibody-drug conjugates that target the protein encoded by *TACSTD2*, known as trophoblast antigen 2 (TROP2), are under active development by several companies. Thus far, Trodelvy (sacituzumab govitecan) has been approved in urothelial cancer [50] and breast cancer (HR+HER2- and TNBC) [51, 52, 53] with active development in NSCLC [54], among other indications. Similarly, Dato-DXd (datopotamab deruxtecan) is actively being pursued in breast cancer [55] and NSCLC [56]. As an oncology target, TROP2 is of interest due to its expression in many solid tumors and limited expression in normal tissues [57]. Moreover, multi-study meta-analyses have shown that over-expression of *TACSTD2* was associated with poor overall survival and reduced disease-free survival [58].

Leveraging our pan-cancer approach, we find that elevated *TACSTD2* expression was predicted pan-cancer with an AUROC of 0.85, in BRCA with an AUROC of 0.63, and in NSCLC with an AUROC of 0.75, as expected. However, the pan-cancer foundation model also suggests predictive power in several additional cancer types, including pancreatic (AUROC: 0.79), stomach (AUROC: 0.89), and thyroid (AROC: 0.73). Previous work has suggested that TROP2 over-expression occurs in these cancer types [59]. Others have also recently reported preclinical evidence of tumor reduction using another TROP2-targeting ADC in xenograft mouse models of pancreatic cancer [60], consistent with our finding.

We further investigated the potential for *TACSTD2* biomarkers by developing specialized, cohort-specific over-expression and signature models in NSCLC and pancreatic cancer. Performance of the resulting model is shown in Figure 5(b). In both cases, specialization improved performance at signature prediction at the cost of some performance in expression prediction. In NSCLC, the AUROC for signature prediction increased from 0.75 to 0.82, and in pancreatic from 0.89 to 0.90. Meanwhile, for over-expression prediction, the AUROC decreased from 0.79 to 0.72 for NSCLC, and 0.79 to 0.78 for pancreatic. In this situation, we could retain the pan-cancer model for predicting over-expression, while deploying the specialized models for amplification signature prediction.

#### Cabozantinib Case Study

A key clinical application of our approach is the ability to use a biomarker to stratify patients into responders and non-responders. Unfortunately, availability of clinical outcomes in our cohorts is limited, especially for targeted therapies, which are often relatively new to clinical practice. To increase the set of hypotheses we could examine, we expanded our evaluation beyond biologics to consider any targeted therapies against our selected targets for which there were a sufficient number of patients (*n >* 30) to properly power our analysis. This resulted in 38 (indication, target) pairs. We then tested for associations between imputed signatures and overall survival (OS) after adjusting for age at diagnosis, age at disease staging, pre-treatment stage, sex, cancer type, metastatic status, number of unique prior therapies, and time from diagnosis to treatment with the therapy of interest.

Our analysis revealed a significant association between our VEGFR2 (*KDR*) amplification signature and OS among cabozantinib-treated patients (hazard ratio [HR]: 0.087; 95% CI, 0.032 to 0.237; Bonferroni adjusted *P* = 7.0 *×* 10*^−^*^5^). The covariate-adjusted Kaplan-Meier curves comparing patients with low versus high VEGFR2 signature scores are presented in Figure 6. Importantly, no outcome data (for cabozantinib or any other drug) were used to inform the design of the amplification signature. For comparison, the HR for the MET signature among the same patient group was 1.39 (95% CI, 0.573 to 3.351; *P* = 0.47). An analysis of measured VEGFR2 expression level also suggested an association with improved OS among patients treated with cabozantinib (HR: 0.727), although the evidence was inconclusive (*P* = 0.10), illustrating the increased power of our imputed signatures. Notably, the VEGFR2 signature (measured or imputed) was not correlated with improved OS in 372 cohort A RCC patients more broadly, suggesting that the clinical benefit is specific to cabozantinib.

**Figure 6:**
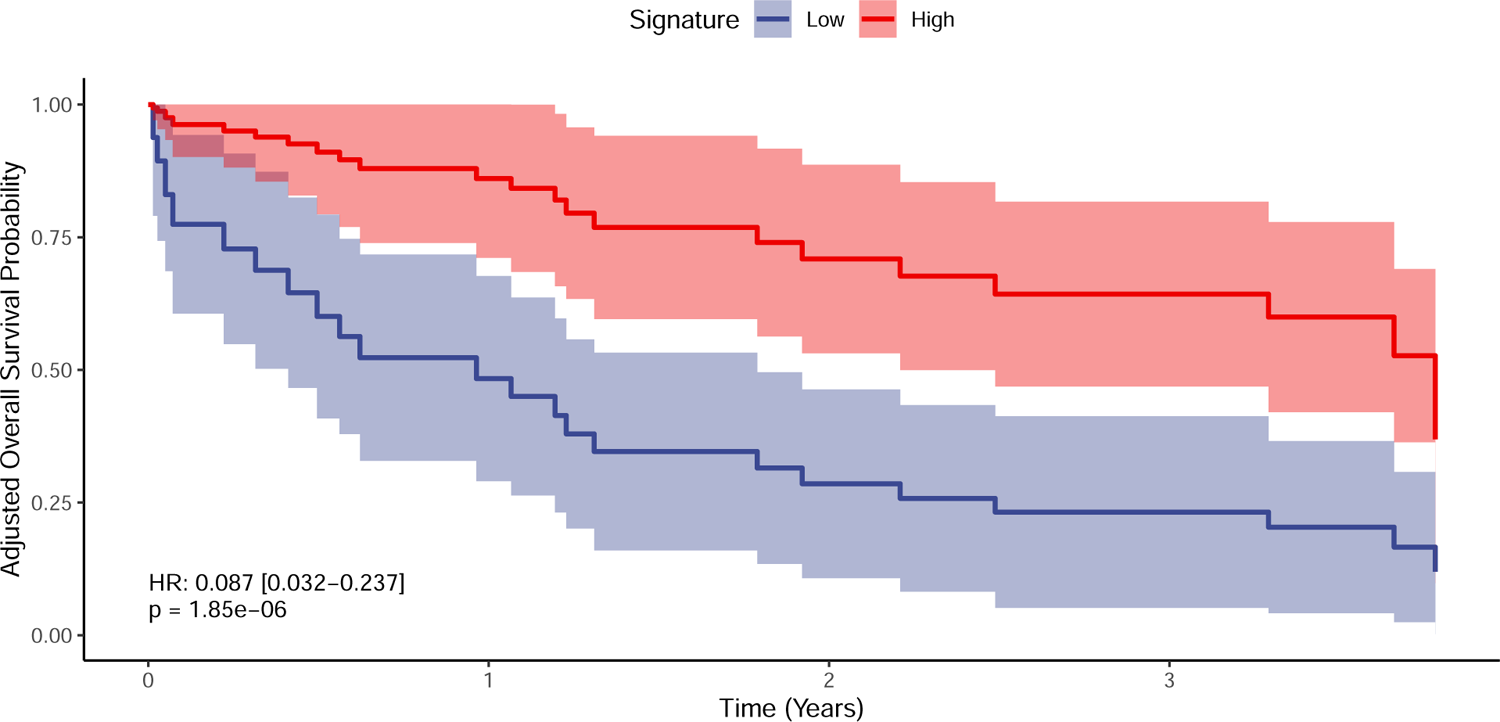
Covariate-adjusted Kaplan-Meier curves for overall survival among patients treated with cabozantinib. Patients were partitioned into two groups (“high” and “low”) on the basis of their VEGFR2 (*KDR*) amplification signature, but without reference to their survival. The reported hazard ratio (HR) and p-value, comparing patients in the low vs. high VEGFR2 signature groups, were estimated via a Cox model, adjusting for clinical covariates. The KM curves were adjusted for covariates using direct standardization.

Cabozantinib is a broad-ranging tyrosine kinase inhibitor (TKI) with activity against MET, RET, AXL, VEGFR2, FLT3, and c-KIT [61], and it has been approved for treatment of renal cell carcinoma (RCC), medullary thyroid cancer, and hepatocellular carcinoma [62]. Of the 31 patients treated with cabozantinib, a majority (22/31) were diagnosed with renal cancer. VEGF-A is a known prognostic marker in metastatic RCC, and high levels of VEGF-A are associated with poorer OS and progression-free survival among patients treated with sunitinib, another TKI [63]. A previous study demonstrated that markers of angiogenesis microvascular density and mast cell density were associated with improved outcomes in metastatic clear cell RCC; however, these did not seem to be predictive of efficacy for cabozantinib compared to everolimus (an mTOR inhibitor) [64]. Despite this finding, given the known relationship of RCC biology with VEGF signaling and the proposed MOA of cabozantinib, the highly significant association between our VEGFR2 signature and OS among cabozantinib-treated patients could be of interest for future biomarker development, as well as providing suggestive evidence for VEGF as a mechanism by which cabozantinib derives efficacy in RCC.

#### Interrogating Spatial Heterogeneity

An important attribute of our model is that it generates biomarker predictions at the resolution of individual tiles. This provides us with the ability to generate spatial gene expression predictions across WSIs. Specifically, the model makes predictions for each biomarker and for each tile, allowing us to create a synthetic annotation on top of the WSI, in which the biomarker predictions are overlaid on each tile within the slide. This capability can be useful in a number of ways. First, it “opens the black box” by providing a human expert the ability to interrogate the process that gave rise to the results. Second, it creates a view on the spatial distribution of multiple biomarkers, providing considerable insight into tumor architecture and intra-tumor heterogenity. Indeed, since these imputations are derived directly from the H&E, this capability supports a form of “label-free” staining across a very large set of molecular readouts.

Figure 7 shows some examples of these synthetic overlays, localizing *HER2* expression in breast cancer and *MET* expression in colorectal cancer. To provide a baseline for these predictions, we also asked our expert pathologist (B.D.) to annotate a random sample of WSIs from patients with and without amplifications while blinded to all model predictions. The fact that increased expression coincides with regions annotated as cancerous aligns with clinical knowledge, and suggests that the model has learned to distinguish between tumor and normal tissue. Importantly, the model has learned this distinction while trained only on bulk, not spatially resolved, expression data. **Supplemental** Figures 20 **and 21** provide an alternate view of these results in which the target expression and amplification signature predictions are juxtaposed.

**Figure 7:**
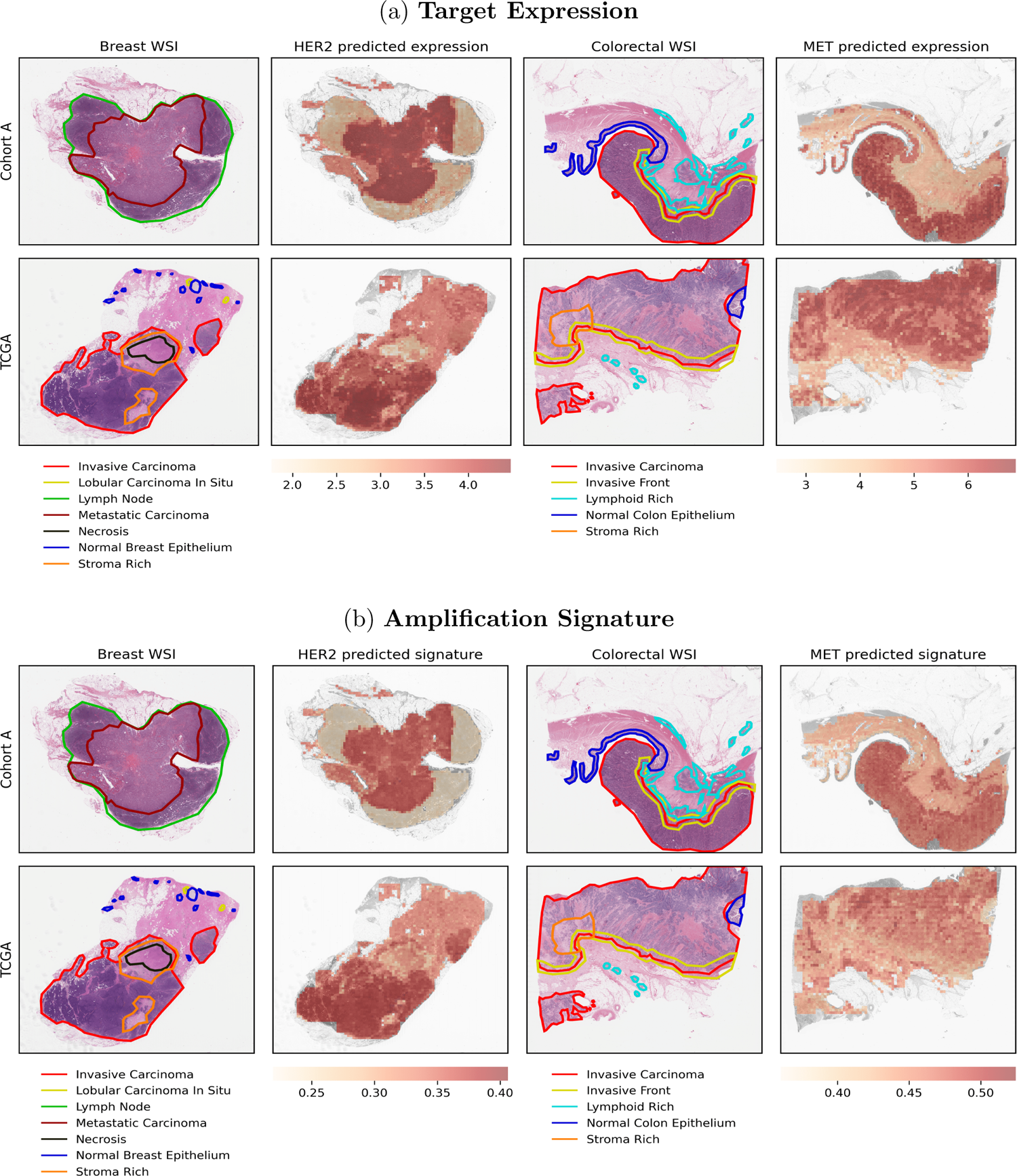
Comparison of expression/signature predictions with expert pathologist annotations. The pathologist was blinded to the predictions. Although the expression/signature models provide tile-level predictions, they were trained only on bulk, not spatially resolved, information. The coincidence of increased expression level and amplification signatures with regions annotated as cancerous therefore provides evidence that the model has learned to differentiate tumor and normal tissue.

Since the molecular labels are all synthetically generated, we can also derive multiple labels for the same image. Figure 8 and **Supplemental** Figure 22 provide examples of co-expression prediction for *HER3* plus *MET* and *TOP1* plus *TOP2A*, respectively. The differences in the spatial expression predictions of these target pairs underscore that the model is learning more nuanced expression information than whether or not a tile falls within a cancerous region of the slide.

**Figure 8:**
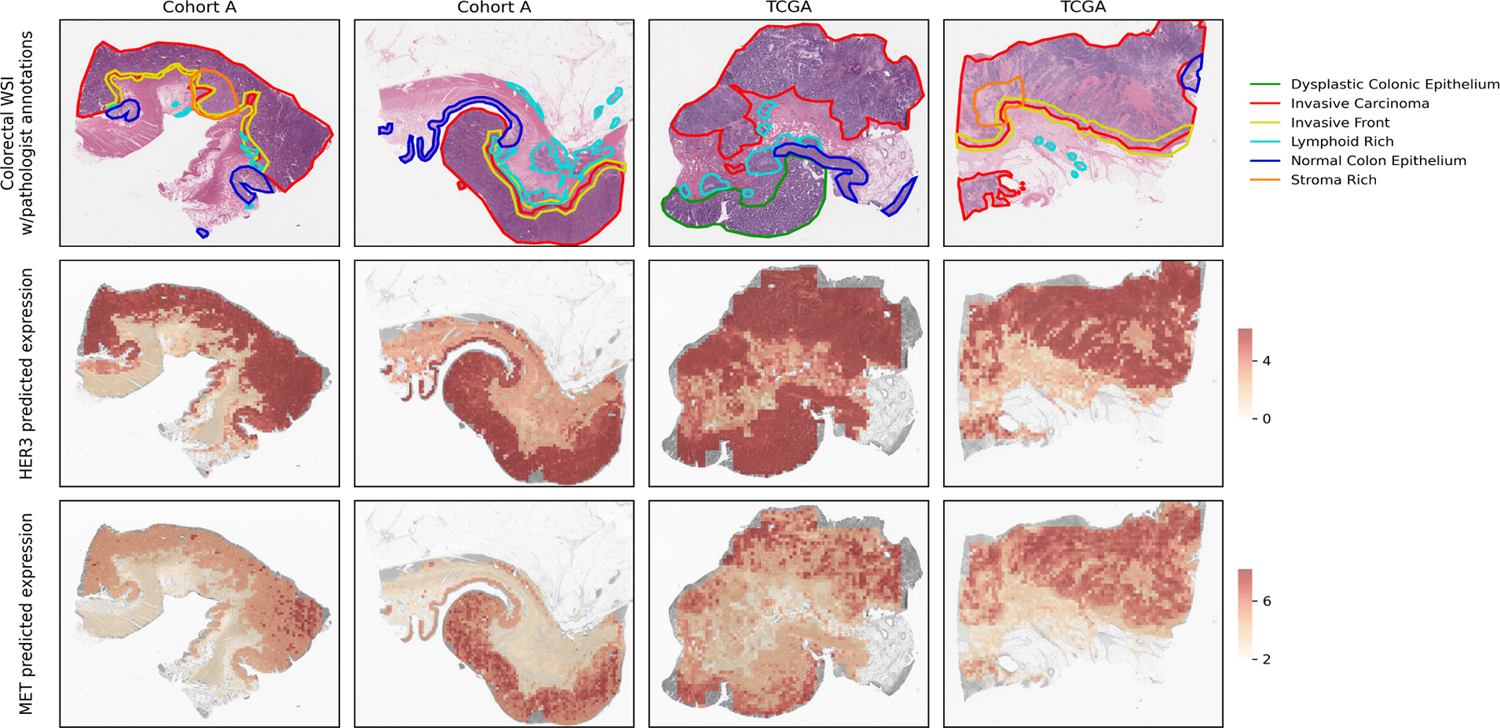
Coexpression of HER3 and MET in colorectal cancer alongside pathologist annotations. The pathologist was blinded to the predictions. Although the expression/signature models provide tile-level predictions, they were trained only on bulk, not spatially resolved, information.

### 3.3 Machine Learning Modeling Insights

#### Transportability Across Cohorts

A critical aspect of machine learning models is the extent to which they generalize outside of the distribution on which they were trained. This generalization is important in assessing the robustness of the approach, i.e., in not overfitting to the specifics of a single data set. It is also critical from a clinical deployment perspective, increasing confidence that the model will behave when applied in a new clinical setting.

**Table 2** presents AUROCs from an experiment where we trained multi-task binary outcome models to predict elevated expression and amplification signatures using data from TCGA only, then evaluated on patients from cohort A only. Results are shown pan-cancer and within breast and colorectal cancer. Stratified results are not presented as the set of cancers available in cohort A only differs from the set available in TCGA + cohort A, and the results would not be comparable with those presented elsewhere. Although significant predictive power is retained, some decrease in performance is always expected when applying a model in a new dataset. Surprisingly, for breast and colorectal cancer, elevated signature prediction improved across datasets, perhaps suggesting these cohorts are more heterogeneous in TCGA than in cohort A.

**Table 2:** Transportability of binary expression and amplification predictions across data sets. Reported are the mean AUROCs across targets. The within dataset results were obtained via cross-validation, training and testing with data from both TCGA and cohort A. The across dataset results were obtained by training on TCGA only then evaluating on cohort A only.

| Biomarker | Cohort | Across Dataset | Within Dataset | Relative Change (%) |
| --- | --- | --- | --- | --- |
| RNA | Pan-cancer | 0.742 | 0.853 | -13.0 |
| RNA | Breast | 0.676 | 0.731 | -7.5 |
| RNA | Colorectal | 0.689 | 0.720 | -4.4 |
| SIG | Pan-cancer | 0.797 | 0.897 | -11.2 |
| SIG | Breast | 0.777 | 0.773 | 0.5 |
| SIG | Colorectal | 0.786 | 0.724 | 8.5 |

#### Machine Learning Architectures

In the course of our work, we experimented with different architectures, summarized in **Table 3**. Panels (a) and (b) explore different ways in which the information across different tiles might be combined. Panel (a) considers whether the model should receive as input separate embeddings for each tile in a patient’s WSI, or the average embedding across tiles. Maintaining separate embeddings for each tile performed better, which essentially provides the model with more, albeit correlated, training examples. Panel (b) considers whether to generate predictions for each tile separately, or to incorporate an attention mechanism, allowing the model to make patient-level predictions while attending to spatially adjacent tiles. While spatial attention did not benefit our models overall, the prediction of certain targets did benefit; for example, in *TLR9* the Spearman correlation between observed and predicted expression increased from 20% to 48%.

**Table 3:**
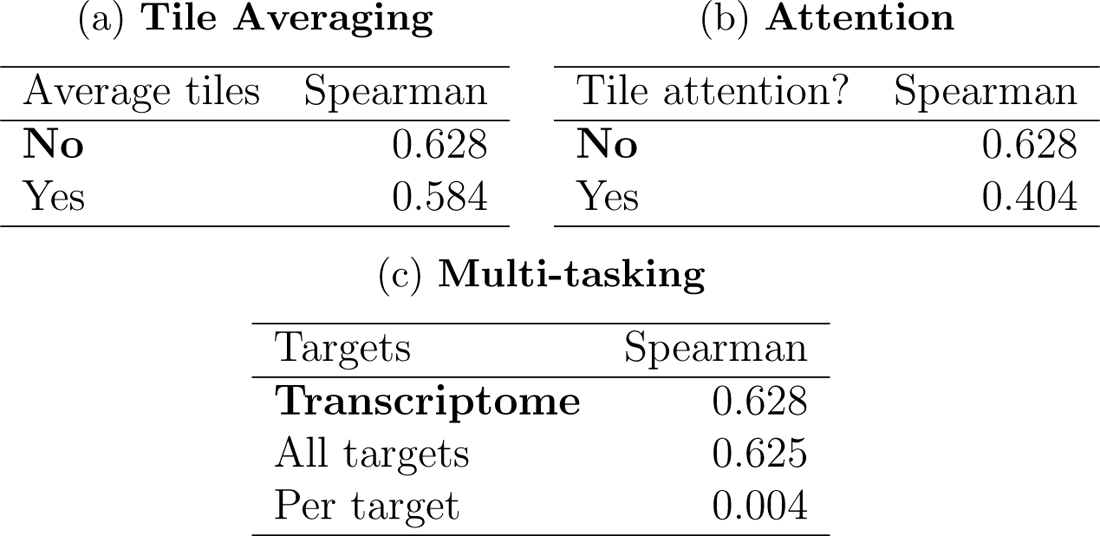
Model architecture experiments. Evaluations were performed pan-cancer via 8-fold cross validation. The reported metric is the mean patient-level correlation across targets. (a) Considers whether the model should receive as input a separate embedding for each tile or the average embedding across tiles. (b) Considers whether to make tile-level predictions independently or to incorporate a spatial attention mechanism. (c) Considers whether to predict expression levels transcriptome-wide then subset the targets of interest, predict expression levels of the targets only, or train separate models for each target.

| (a) **Tile Averaging** |  | (b) **Attention** |  |
| --- | --- | --- | --- |
| Average tiles | Spearman | Tile attention? | Spearman |
| **No** | 0.628 | **No** | 0.628 |
| Yes | 0.584 | Yes | 0.404 |

| (c) **Multi-tasking** |  |
| --- | --- |
| Targets | Spearman |
| **Transcriptome** | 0.628 |
| All targets | 0.625 |
| Per target | 0.004 |

Panel (c) considers the value of training across multiple biomarker tasks. Specifically, we compare an approach where we train separate models to predict each target, train a single model to simultaneously predict all targets, or train a single model to predict expression transcriptome-wide then subset the targets of interest. The last strategy performed best, and both multi-task strategies substantially outperformed the individual (single-task) strategy for our model architecture.

## 4 Discussion

At the core of our work is a demonstration of the ability of machine learning models, using standard H&E images as input, to impute multiple types of molecular covariates of potential clinical significance. Specifically, we showed high quality imputation of both copy number amplifications (CNAs) and transcriptome-wide RNA-seq. We also defined, for each of our target genes, a derived signature that aims to capture the transcriptional consequences of a CNA in that gene; this signature provides a quantitative biomarker for CNA “phenocopies” [65]. Our ML models achieve strong predictive accuracies across all three types of biomarkers. For both RNA-seq and amplification signatures, we achieved good predictions across the spectrum of expression levels, as measured by correlation coefficients. And across all three modalities, we demonstrated high accuracy in identifying individuals whose biomarker levels are elevated, corresponding to a predicted amplification or overexpression event.

### Machine Learning Lessons

A key factor in the strong performance of our ML models is that they exploit commonalities of cancer mechanisms across cancer types, as well as different genes and mutations, as they manifest both in the H&E slides and in the molecular readouts. In this regard, our work stands in contrast to the standard approach in digital pathology, where a model is usually trained separately for each biomarker prediction task. For example, in the work of Arsalan *et al* [16], over 13,000 distinct models are trained, one for each cancer, biomarker, and fold. The recently published H&E foundation models take a significant step forward, training a single model that provides a uniform, pan-cancer featurization. Indeed, Chen *et al* [9] show that these foundation models are able to provide at-or-beyond state-of-the-art performance across a very diverse range of tasks. However, they typically use single task-specific heads that do not attempt to leverage shared mechanisms across different prediction tasks, such as different biomarkers. In our work, we showed that the predictive performance of our models increases significantly as we move from predicting a single molecular readout to a multi-task prediction across all of our target genes to a transcriptome-wide prediction. A broad comparison to the results of Arslan *et al* is challenging due to the very limited overlap in biomarkers between their work and ours. However, for CDK4, which is the only shared RNA biomarker, they report at AUROC of around 0.72 (averaged across 3 cancer types) [16], whereas we obtain an AUROC of 0.84 pan-cancer; of 0.75 in a stratified analysis, averaged across all cancer types; and of 0.77 when filtering to 4 relevant cancer types (specifically, breast, colorectal, lung, and pancreatic).

The potential downside of our pan-cancer, pan-biomarker approach is that it may not maximally learn to recognize features that capture tumor- or biomarker-specific variation and instead may focus on learning features that generalize across tasks. As we showed, this limitation can be addressed by refining our base model to a specific prediction task; this can be a single cancer, a single biomarker, or a combination of both. As we showed in our MET case study, these fine-tuned models can provide improved performance for a specific prediction task. A similar process could be applied to fine-tune a biomarker predictive model to a treatment-response data set for a drug with a relevant MOA, shifting the model towards better predicting patient response. Because of the extensive pre-training of the model, such fine-tuning might be feasible even from the small-scale cohorts available in phase 1/2 clinical trials.

Our models achieve considerably higher performance at predicting RNA and even higher performance at predicting amplification signatures. We believe that this higher performance derives from several sources. First, quantitative traits offer enhanced statistical power over binary or ordinal traits such as CNA, since continuous data capture more granular phenotypic variation, and provide meaningful information across the entire set of individuals, significantly increasing the effective sample size. Second, multiple studies have shown that copy number amplification is only one mechanism by which clinically relevant activation of a gene or pathway might be achieved, and other mechanisms might converge on the same pathway, resulting in the same phenotypic consequences. Our use of alternative genome-wide signatures captures a broader range of these “CNA phenocopy” mechanisms [65], and avoids creating artificial and biologically meaningless distinctions in the training set, which serve to confuse the ML model. Moreover, there are indications [65, 66] that patients lacking a mutation in a particular target but with a transcriptional pattern concordant with that mutation may benefit from the same class of treatments as a patient harboring a genuine amplification.

One important extension of our work is to train the models to predict protein levels, since proteins are the direct therapeutic targets of the majority of drugs (including those that we focused on in our study). Notably, Arsalan *et al* [16] achieve a higher predictive accuracy on protein biomarkers than on RNA. Given our results, we believe that a multi-task approach would be highly beneficial in this context as well. Moreover, the generality of our framework would allow multi-task learning on multiple biomarker types in the same model, enabling information transfer across CNA, RNA, protein, and more.

Recognizing the importance of spatial heterogeneity of tumors, we also considered several machine learning approaches for integrating across different regions in the image: averaging the featurization of different tiles prior to making predictions; using a multiple-instance learning (MIL) approach, where the model is equipped with an attention mechanism to select relevant subsets of the image; and making individual tile-level predictions then averaging them. With a few exceptions, the latter approach provided considerably better performance. We believe this to be the case because the models are trained to predict bulk transcription levels, which is concordant with a model that predicts these profiles per tile, and then averages the results. By comparison, averaging before making predictions attenuates the signal and loses resolution, whereas the MIL approach selects predictions from one region, which may not fully capture the signal from across the tumor. Previous work [37] has demonstrated the benefits of MIL in other contexts; it would be interesting to understand which tasks lend themselves better to MIL versus other aggregation approaches.

Despite being trained on bulk transcription levels, our models learn spatial variation in expression levels and amplification signatures that strongly correlate with, but do not simply recapitulate, the difference between tumor and normal tissue. This is seen from the variation across targets in our spatial co-expression examples. Comiter *et al* [67] recently developed ML models that produce spatially resolved ‘omics data from H&E, using a training set of aligned H&E and scRNA-seq (versus bulk) data, and showed reasonable spatial correlation, at the tile level, with both pathologist scores and measured spatial expression. In ongoing work, we are collecting spatial expression data, which would allow a more quantitative assessment of our spatial predictions and even training models to directly predict measured spatial ‘omics. We expect this to further increase the accuracy and potential clinical utility of our predictions.

### Potential Impact on Preclinical Development

Our approach enables us to derive full molecular profiles from routinely collected histopathology images, defining a “semi-synthetic” cohort where imputed molecular data, inferred from real H&E, complements other measured covariates, including patient demographics, medical histories, treatments, and clinical outcomes. Given the abundance of cohorts that comprise H&E alongside these other covariates, one can produce a very large semi-synthetic cohort that is highly powered for a broad set of exploratory analyses. Specifically, we can explore and even construct diverse multi-modal biomarkers, assess which are well-predicted, and estimate associations with clinically relevant covariates (such as CNAs, survival, or treatment response). Indeed, our CNA signature biomarkers were derived directly from the imputed RNA profiles, allowing us to substitute a hard-to-estimate (and potentially limited) biomarker with one that can be estimated much more robustly; other signatures, which combine RNA measurements in different ways, can be defined and evaluated similarly. As discussed above, these predictions could then be fine-tuned using data from phase 1/2 clinical trials. Overall, our approach provides simultaneous imputations across multiple molecular modalities and numerous targets, enabling a quantitative assessment of multiple “response signatures” and the data-driven selection of a biomarker that could be deployed in the design and conduct of phase 3 studies and ultimately in personalized medicine.

In addition to identifying biomarkers for a given target within a select tumor type, our approach also enables the identification of potential new therapeutic opportunities. Specifically, our pan-cancer results demonstrate an ability to accurately impute expression levels across multiple cancers from very diverse tissues of origin. These predictions can help highlight cancers where a cancer target is significantly expressed, at a level that might be therapeutically relevant (as compared with other cancers where that MOA is deployed). This could suggest new opportunities to expand the set of indications for a given targeted drug. While these insights could potentially be derived from molecular data collected across tumor types, such data are not regularly collected as part of the standard of care, making it difficult to detect those opportunities, especially for rare cancers and/or smaller patient subpopulations. In cases where clinical outcomes in response to treatment are available, we can also test for associations between a signature and these outcomes. As demonstrated in our preliminary analysis on cabozantinib response, these associations could potentially inform our understanding of which aspect of the drug’s MOA is driving efficacy, and hence suggest potential avenues for generating improved chemical matter.

### Potential Clinical Impact

We selected targets where existing drugs derive tumor-killing efficacy directly from the abundance of the proteins; therefore, there is reason to believe that variation in target abundance will have an effect on therapeutic response. However, our biomarkers do not directly correspond to protein abundance, potentially limiting their predictive power in some cases. There are also other factors that might create a discrepancy between target abundance and therapeutic benefit. The ultimate test of the value of our biomarkers is measuring how well they predict outcomes among patients treated with the corresponding drugs. Due to very limited availability of objective clinical outcome data for these standard-of-care drugs, we were only able to perform this assessment for a limited number of targeted therapies, as discussed above. The preliminary results from the cabozantinib case study demonstrate the potential for a machine-learning defined signature to be predictive of superior clinical outcomes for a specific targeted therapy without requiring training on any response or outcome data for that drug.

Scaling this analysis to a broader set of signature-therapy combinations is a critical direction for future research and an ongoing focus of our work. If successful, our approach could enable the use of the H&E images that are ubiquitously collected to identify patients that are likely to benefit from targeted therapies. This capability can be deployed in a variety of ways. Most immediately, it can be used as a rapid triage step for suggesting a set of therapeutic interventions that might be relevant to a patient; this step could be followed by the deployment of other, more standard, biomarker assay(s) such as genetic sequencing or IHC, to verify that the patient is indeed eligible for the drug, given the currently approved label. Importantly, such ML-based H&E biomarkers could be deployed rapidly, across geographies, and without specialized equipment or reagents beyond H&E staining and scanning, making them accessible on a broad scale. In the longer term, however, our work opens the opportunity to directly use an individual patient’s H&E-derived biomarkers to identify and prescribe therapeutic interventions. Unlike most other biomarkers, which generally focus on one or two molecular measurements, our H&E biomarkers rely on the full context of whole-slide images, which provide a broad, detailed, multi-scale phenotype. As such, it is possible that they may detect more diffuse slide-level evidence that better captures coherent groups of patients that may have similar outcomes on treatment. This analysis might help identify patients who are unlikely to benefit, enabling a clinician to suggest a different course of treatment. On the other side, new patients might be identified. Indeed, the sets of patients at the higher end (95th percentile) of our RNA and amplification signature biomarkers are considerably larger than those defined by CNAs directly; thus, our biomarkers could help expand the population of patients who might benefit from a drug. Notably, this approach is generalizable across a broad set of targeted therapies.

## Conclusion

Overall, our work highlights the latent potential in the enormous amounts of H&E images that are regularly collected as part of standard patient care. We have shown that a surprisingly large set of molecular measurements can be robustly imputed from these H&E images. The scale of the resulting semi-synthetic cohorts, where H&E and clinical covariate data are complemented with imputed molecular measurements, generates massive statistical power that can power novel insights in preclinical discovery. The power and flexibility of these digital biomarkers allows us to define specialized signatures that might better capture the patient population likely to benefit from a drug. Moreover, digital H&E-derived biomarkers do not require unique reagents or advanced assays. The ubiquity of H&E data, and the increasing digitization of this modality, therefore unlock the opportunity to rapidly develop novel biomarkers and deploy them to broad patient populations around the world.

## Data Availability

Data produced in the present study are available upon reasonable request to the authors.

## References

[1] A Marabelle, DT Le, PA Ascierto, et al. Efficacy of pembrolizumab in patients with noncolorectal high microsatellite instability/mismatch repair-deficient cancer: Results from the phase ii keynote-158 study. J Clin Oncol, 38(1):1 – 10, 2020.

[2] TK Choueiri, T Powles, L Albiges, et al. Cabozantinib plus nivolumab and ipilimumab in renal-cell carcinoma. N Engl J Med, 388(19):1767 – 1778, 2023.

[3] JL Parker, SS Kuzulugil, K Pereverzev, et al. Does biomarker use in oncology improve clinical trial failure risk? a large-scale analysis. Cancer Medicine, 10(6):1955 – 1963, 2021.

[4] L Mohamed, S Manjrekar, DP Ng, et al. The effect of biomarker use on the speed and duration of clinical trials for cancer drugs. Oncologist, 27(10):849 – 856, 2022.

[5] MKK Niazi, AV Parwani, and M Gurcan. Digital pathology and artificial intelligence. Lancet Oncology, 20(5):e253 – e261, 2019.

[6] MG Hanna, O Ardon, VE Reuter, et al. Integrating digital pathology into clinical practice. Modern Pathology, 35(2):152 – 164, 2022.

[7] V Baxi, R Edwards, M Montalto, and S Saha. Digital pathology and artificial intelligence in translational medicine and clinical practice. Modern Pathology, 35(1):23 – 32, 2022.

[8] H Dawson. Digital pathology – rising to the challenge. Frontiers in Medicine, 9:888896, 2022.

[9] RJ Chen, T Ding, MY Lu, et al. A general-purpose self-supervised model for computational pathology. arXiv, 2023.

[10] P Raciti, J Sue, R Ceballos, et al. Novel artificial intelligence system increases the detection of prostate cancer in whole slide images of core needle biopsies. Modern Pathology, 33(10):2058 – 2066, 2020.

[11] L da Silva, EM Pereira, PG Salles, et al. Independent real-world application of a clinical-grade automated prostate cancer detection system. J Pathol, 254(2):147 – 158, 2021.

[12] S Perincheri, AW Levi, R Celli, et al. An independent assessment of an artificial intelligence system for prostate cancer detection shows strong diagnostic accuracy. Modern Pathology, 34(8):1588 – 1595, 2021.

[13] C Saillard, R Dubois, O Tchita, et al. Validation of msintuit as an ai-based pre-screening tool for msi detection from colorectal cancer histology slides. Nature Communications, 14(1):6695, 2023.

[14] OL Saldanha, CML Loeffler, JM Niehues, et al. Self-supervised attention-based deep learning for pan-cancer mutation prediction from histopathology. NPJ Precision Oncology, 7(1):35, 2023.

[15] J Anaya, JW Sidhom, F Mahmood, and AS Baras. Multiple-instance learning of somatic mutations for the classification of tumour type and the prediction of microsatellite status. Nature Biomedical Engineering, 2023. Online ahead of print.

[16] S Arslan, D Mehrotra, J Schmidt, et al. Deep learning can predict multi-omic biomarkers from routine pathology images: A systematic large-scale study. bioRxiv, 2022.

[17] E Vorontsov, A Bozkurt, A Casson, et al. Virchow: A million-slide digital pathology foundation model. arXiv, 2023.

[18] S Pai, D Bontempi, I Hadzic, et al. Foundation models for quantitative biomarker discovery in cancer imaging. medRxiv, 2023.

[19] M Moor, O Banerjee, ZSH Abad, et al. Foundation models for generalist medical artificial intelligence. Nautre, 616(7956):259 – 265, 2023.

[20] Cancer Genome Atlas Research Network, JN Weinstein, EA Collisson, et al. The cancer genome atlas pan-cancer analysis project. Nature Genetics, 45(10):1113 – 1120, 2013.

[21] CH Mermel, SE Schumacher, B Hill, et al. Gistic2.0 facilitates sensitive and confident localization of the targets of focal somatic copy-number alteration in human cancers. Genome Biology, 12(4):R41, 2011.

[22] F Favero, T Joshi, AM Marquard, et al. Sequenza: allele-specific copy number and mutation profiles from tumor sequencing data. Annals of Oncology, 26(1):64 – 70, 2015.

[23] Genomic data commons data portal, 2023. Accessed: November 1st, 2023.

[24] CW Law, Y Chen, W Shi, and GK Smyth. voom: Precision weights unlock linear model analysis tools for rna-seq read counts. Genome Biology, 15(2):R29, 2014.

[25] R Core Team. R: A Language and Environment for Statistical Computing. R Foundation for Statistical Computing, Vienna, Austria, 2021.

[26] MD Robinson, DJ McCarthy, and GK Smyth. edger: a bioconductor package for differential expression analysis of digital gene expression dat. Bioinformatics, 26(1):139 – 140, 2010.

[27] M Macenko, M Niethammer, JS Marron, et al. A method for normalizing histology slides for quantitative analysis. IEEE International Symposium on Biomedical Imaging: From Nano to Macro, pages 1107 – 1110, 2009.

[28] A Dosovitskiy, L Beyer, A Kolesnikov, et al. An image is worth 16×16 words: Transformers for image recognition at scale. In International Conference on Learning Representations, 2021.

[29] M Caron, H Touvron, I Misra, et al. Emerging properties in self-supervised vision transformers. arXiv, 2021.

[30] Z Shao, H Bian, Y Chen, et al. TransMIL: Transformer based correlated multiple instance learning for whole slide image classification. In A Beygelzimer, Y Dauphin, P Liang, and J Wortman Vaughan, editors, Advances in Neural Information Processing Systems, 2021.

[31] DP Kingma and J Ba. Adam: A method for stochastic optimization. *arXiv*, 2014.

[32] C Szegedy, V Vanhoucke, S Ioffe, J Shlens, and Z Wojna. Rethinking the inception architecture for computer vision. In 2016 IEEE Conference on Computer Vision and Pattern Recognition (CVPR), pages 2818 – 2826, 2016.

[33] J Liu, T Lichtenberg, KA Hoadley, et al. An integrated tcga pan-cancer clinical data resource to drive high-quality survival outcome analytics. Cell, 173(2):400 – 416, 2018.

[34] DR Cox. Regression models and life tables. JRSSB, 34(2):187 – 220, 1972.

[35] IM Chang, R Gelman, and M Pagano. Corrected group prognostic curves and summary statistics. Journal of Chronic Diseases, 35(8):669 – 674, 1982.

[36] RW Makuch. Adjusted survival curve estimation using covariates. Journal of Chronic Diseases, 35(6):437 – 443, 1982.

[37] X Shao, N Lv, J Liao, et al. Copy number variation is highly correlated with differential gene expression: a pan-cancer study. BMC Medical Genetics, 20(1):175, 2019.

[38] J Ko, J Jung, ST Kim, et al. Met gene alterations predict poor survival following chemotherapy in patients with advanced cancer. Pathology and Oncology Research, 28:1610697, 2022.

[39] S Park, YL Choi, CO Sung, et al. High met copy number and met overexpression: poor outcome in non-small cell lung cancer patients. Histol Histopathol, 27(2):197 – 207, 2012.

[40] S Al-Saad, E Richardsen, TK Kilvaer, et al. The impact of met, igf-1, igf1r expression and egfr mutations on survival of patients with non-small-cell lung cancer. PLoS One, 12(7):e0181527, 2017.

[41] DR Camidge, J Bar, H Horinouchi, et al. Telisotuzumab vedotin (teliso-v) monotherapy in patients (pts) with previously treated c-met–overexpressing (oe) advanced non-small cell lung cancer (nsclc). Journal of Clinical Oncology, 40(16 suppl):9016–9016, 2022.

[42] R Guo, LD Berry, DL Aisner, et al. Met ihc is a poor screen for met amplification or met exon 14 mutations in lung adenocarcinomas: Data from a tri-institutional cohort of the lung cancer mutation consortium. Journal of Thoracic Oncology, 14(9):1666 – 1671, 2019.

[43] L Landi, G Minuti, A D’Incecco, J Salvini, and F Cappuzzo. Met overexpression and gene amplification in nsclc: a clinical perspective. Lung Cancer (Auckl), 4:15 – 25, 2013.

[44] K Ingale, SH Hong, JSK Bell, et al. Prediction of met overexpression in non-small cell lung adenocarcinomas from hematoxylin and eosin images. *arXiv*, 2023. preprint.

[45] JK Lennerz, EL Kwak, A Ackerman, et al. Met amplification identifies a small and aggressive subgroup of esophagogastric adenocarcinoma with evidence of responsiveness to crizotinib. Journal of Clinical Oncology, 29(36):4803 – 4810, 2011.

[46] YH Xie, YX Chen, and JY Fang. Comprehensive review of targeted therapy for colorectal cancer. Signal Transduct Target Ther, 5(1):22, 2020.

[47] MFD Renzo, M Olivero, A Giacomini, et al. Overexpression and amplification of the met/hgf receptor gene during the progression of colorectal cancer. Clin Cancer Res, 1(2):147 – 154, 1995.

[48] WS El-Deiry, N Vijayvergia, J Xiu, et al. Molecular profiling of 6,892 colorectal cancer samples suggests different possible treatment options specific to metastatic sites. Cancer Biol Ther, 16(12):1726 – 1737, 2015.

[49] SJ Lee, J Lee, SH Park, et al. c-met overexpression in colorectal cancer: A poor prognostic factor for survival. Clin Colorectal Cancer, 17(3):165 – 169, 2018.

[50] RL Wong, LA Ferris, OA Do, et al. Efficacy of platinum rechallenge in metastatic urothelial carcinoma after previous platinum-based chemotherapy for metastatic disease. Oncologist, 26(12):1026 – 1034, 2021.

[51] A Bardia, SA Hurvitz, SM Tolaney, et al. Sacituzumab govitecan in metastatic triplenegative breast cancer. N Engl J Med, 384:1529 – 1541, 2021.

[52] HS Rugo, A Bardia, F Marme, et al. Overall survival with sacituzumab govitecan in hormone receptor-positive and human epidermal growth factor receptor 2-negative metastatic breast cancer (tropics-02): a randomised, open-label, multicentre, phase 3 trial. Lancet, 402(10411):1423 – 1433, 2023.

[53] D Kwapisz. Sacituzumab govitecan-hziy in breast cancer. Am J Clin Oncol, 45(7):279 – 285, 2022.

[54] MC Garassino, D Reznick, SY Liu, et al. Evoke-01: A phase 3 study of sacituzumab govitecan (sg) versus docetaxel in patients with non–small cell lung cancer (nsclc) progressing on or after platinum-based chemotherapy and checkpoint inhibitors. Journal of Clinical Oncology, 40(16 suppl):TPS9149–TPS9149, 2022.

[55] A Bardia, K Jhaveri, S Im, et al. Datopotamab deruxtecan (dato-dxd) vs chemotherapy in previously-treated inoperable or metastatic hormone receptor-positive, her2-negative (hr+/her2–) breast cancer (bc): Primary results from the randomised phase iii tropion-breast01 trial. Annals of Oncology, 34(suppl 2):S1254 – S1335, 2023.

[56] M Ahn, AE Lisberg, L Paz-Ares, et al. Datopotamab deruxtecan (dato-dxd) vs docetaxel in previously treated advanced/metastatic (adv/met) non-small cell lung cancer (nsclc): Results of the randomized phase iii study tropion-lung01. Annals of Oncology, 34(suppl 2):S1254 – S1335, 2023.

[57] DM Goldenberg, R Stein, and RM Sharkey. The emergence of trophoblast cell-surface antigen 2 (trop-2) as a novel cancer target. Oncotarget, 9(48):28989 – 29006, 2018.

[58] P Xu, Y Zhao, K Liu, et al. Prognostic role and clinical significance of trophoblast cell surface antigen 2 in various carcinomas. Cancer Manag Res, 9:821 – 837, 2017.

[59] A Shvartsur and B Bonavida. Trop2 and its overexpression in cancers: regulation and clinical/therapeutic implications. Genes Cancer, 6(3):84 – 105, 2015.

[60] LP Sun, WQ Bai, DD Zhou, et al. himb1636-mmae, a novel trop2-targeting antibodydrug conjugate exerting potent antitumor efficacy in pancreatic cancer. J Med Chem, 66(21):14700 – 14715, 2023.

[61] C Grullich. Cabozantinib: Multi-kinase inhibitor of met, axl, ret, and vegfr2. Recent Results Cancer Res, 211:67 – 75, 2018.

[62] P Maroto, C Porta, J Capdevila, et al. Cabozantinib for the treatment of solid tumors: a systematic review. Ther Adv Med Oncol, 14:17588359221107112, 2022.

[63] Z Guillaume, M Auvray, Y Vano, et al. Renal carcinoma and angiogenesis: Therapeutic target and biomarkers of response in current therapies. Cancers (Basel), 14(24):6167, 2022.

[64] T Denize, S Farah, A Cimadamore, et al. Biomarkers of angiogenesis and clinical outcomes to cabozantinib and everolimus in patients with metastatic renal cell carcinoma from the phase iii meteor trial. Clin Cancer Res, 28(4):748 – 755, 2022.

[65] H Bakhtiar, KT Helzer, Y Park, et al. Identification of phenocopies improves prediction of targeted therapy response over dna mutations alone. NPJ Genomic Medicine, 7(1):58, 2022.

[66] J de Bono, J Mateo, K Fizazi, et al. Olaparib for metastatic castration-resistant prostate cancer. New England Journal of Medicine, 382(22):2091 – 2102, 2020.

[67] C Comiter, ED Vaishnav, M Ciampricotti, et al. Inference of single cell profiles from histology stains with the single-cell omics from histology analysis framework (schaf). *bioRxiv*, 2023. preprint.

