## Supplemental for "Machine learning enabled prediction of digital biomarkers from whole slide histopathology images"

### 1 Supplemental Results

#### 1.1 Copy Number Amplifications

##### 1.1.1 Amplification prevalence

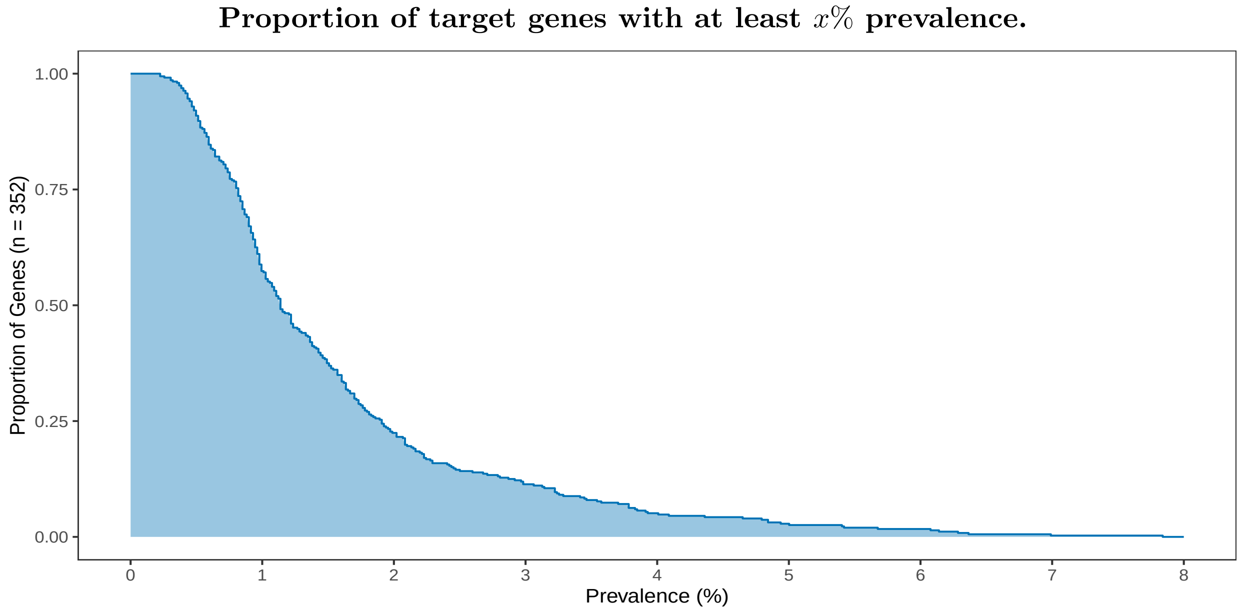

Figure 1: **Distribution of amplification prevalence ($n = 14,007$).**

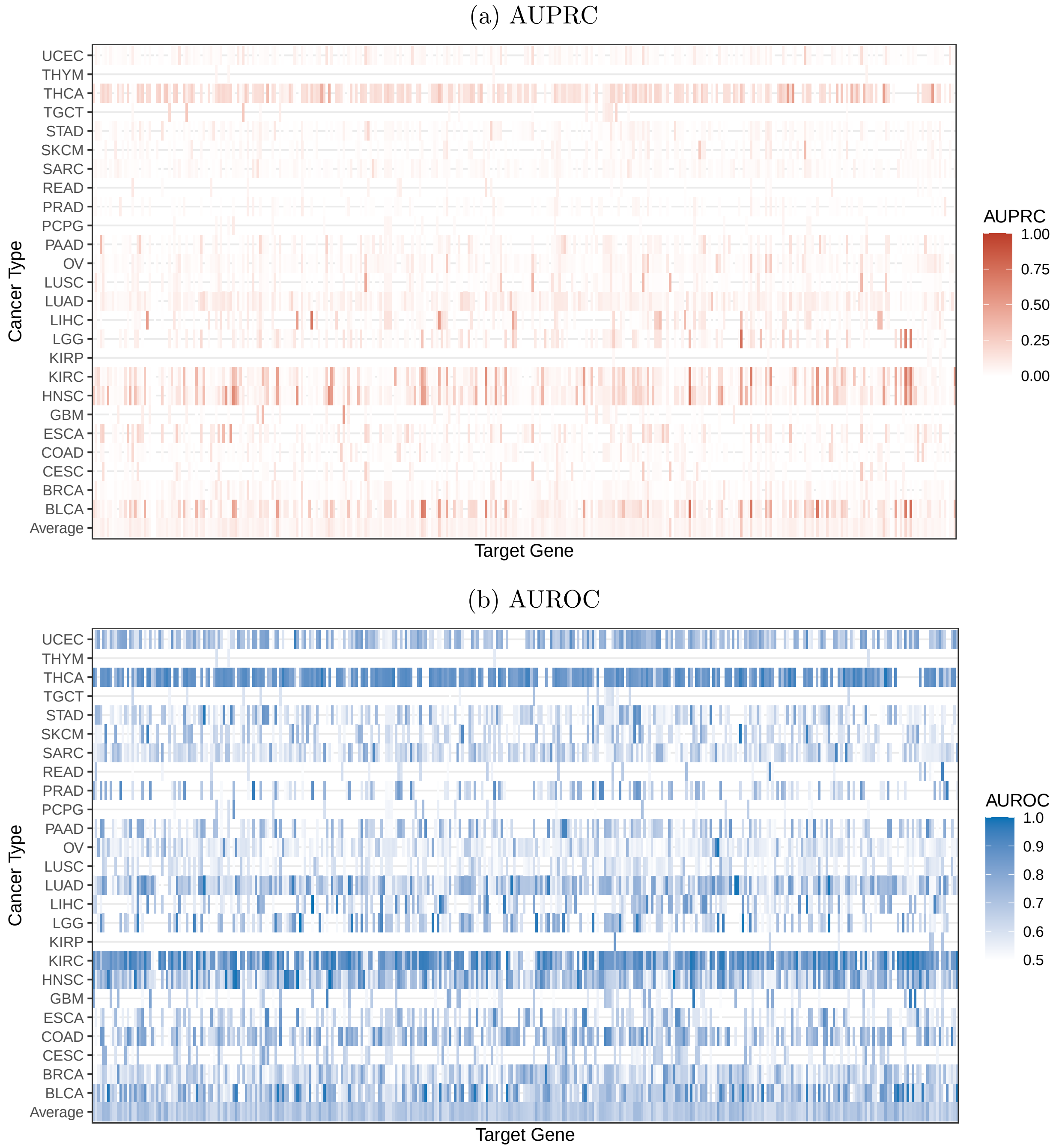

Figure 2: **Prediction of copy number amplifications from digital histopathology, stratified by cancer type.** Performance is evaluated at the patient level in a held-out evaluation set and averaged across 8 cross-validation folds. Metrics are only presented where at least 3 patients within a cancer type harbored a mutation in the target gene.

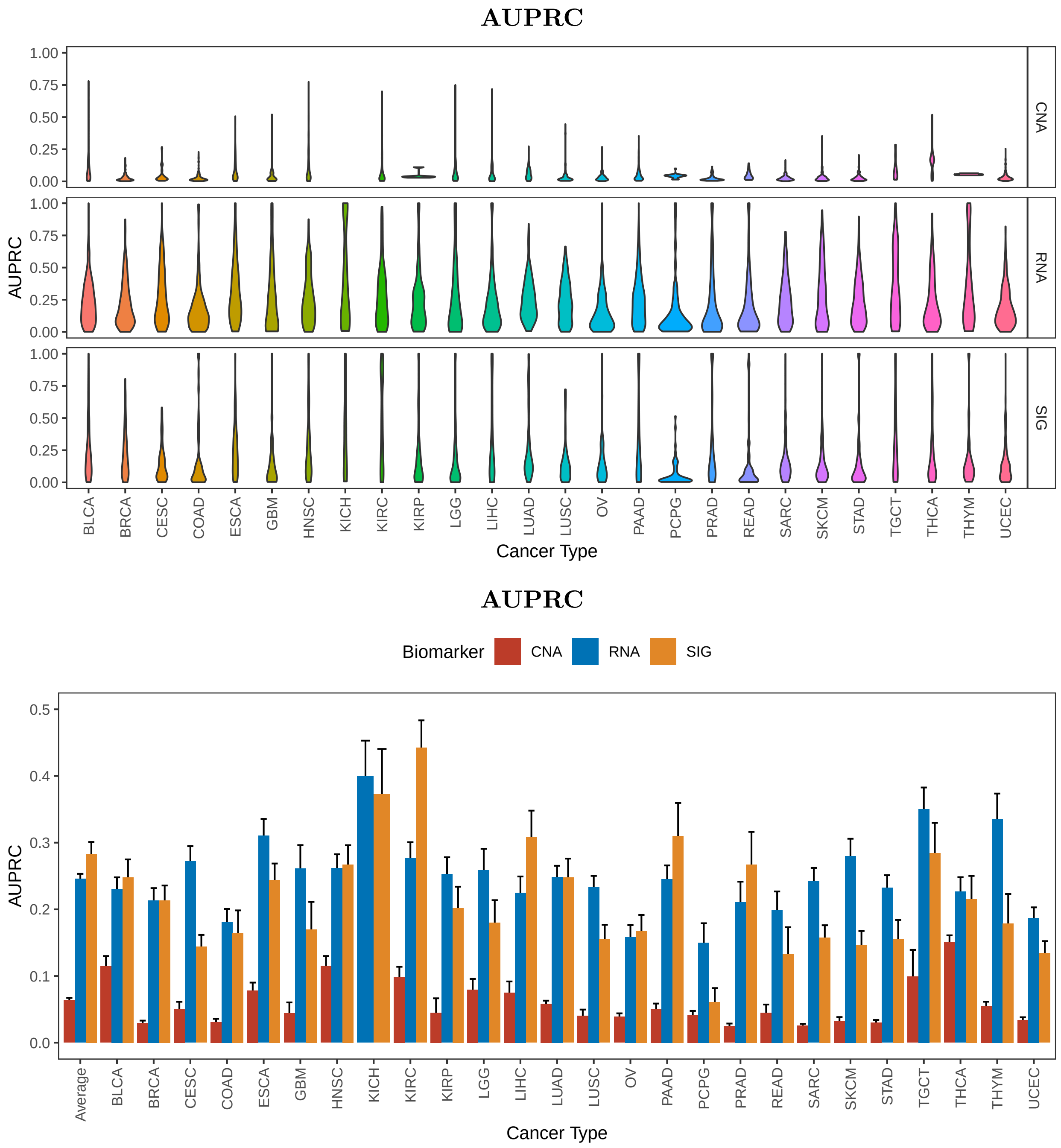

Figure 3: **Cross-modality comparison of binary digital biomarker prediction quality, stratified by cancer-type.** Performance is evaluated at the patient level in a held-out evaluation set and averaged across 8 cross-validation folds. Metrics are calculated separately in each cancer type with $\geq$ 100 patients. The distributions are shown across up to 352 target genes. For CNA, the task was to predict whether the patient harbored an amplification. For target expression (RNA) and the amplification signature (SIG), the task was to predict whether the patient's expression/signature level exceeded the 95th percentile.

##### 1.1.2 Association with expression

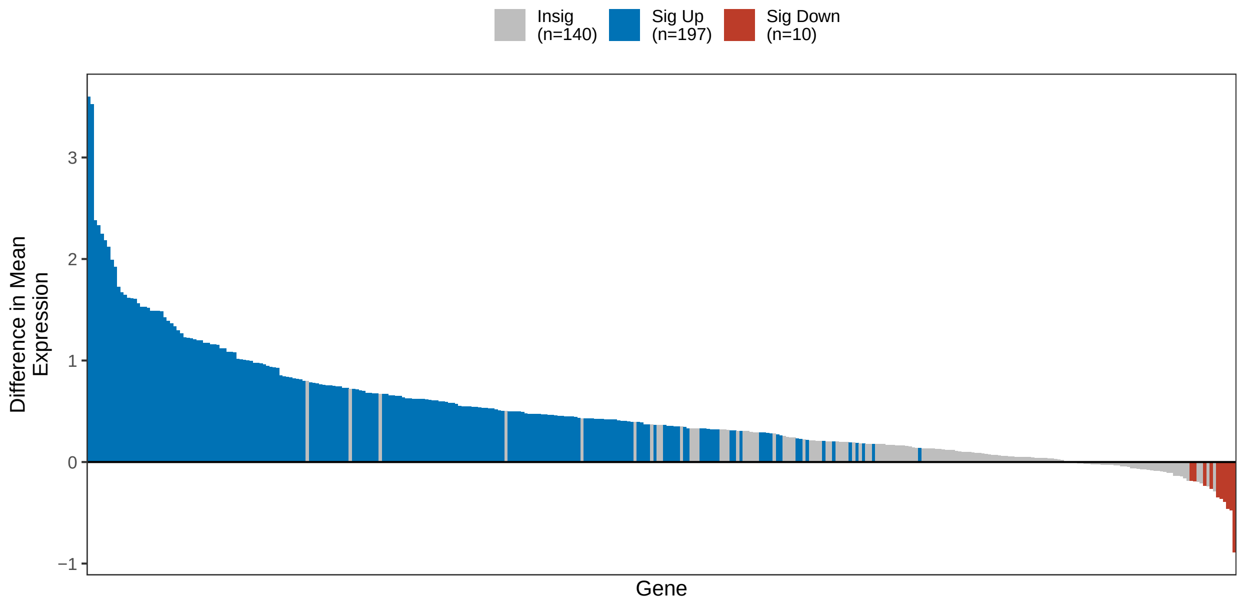

Figure 4: **Difference in expression by gene amplification status.** Genes where the difference in mean expression was insignificant are colored in gray. Among differentially expressed genes, those up-regulated are colored in blue, and those down-regulated in red.

#### 1.2 Target Expression

##### 1.2.1 Expression and signature prediction

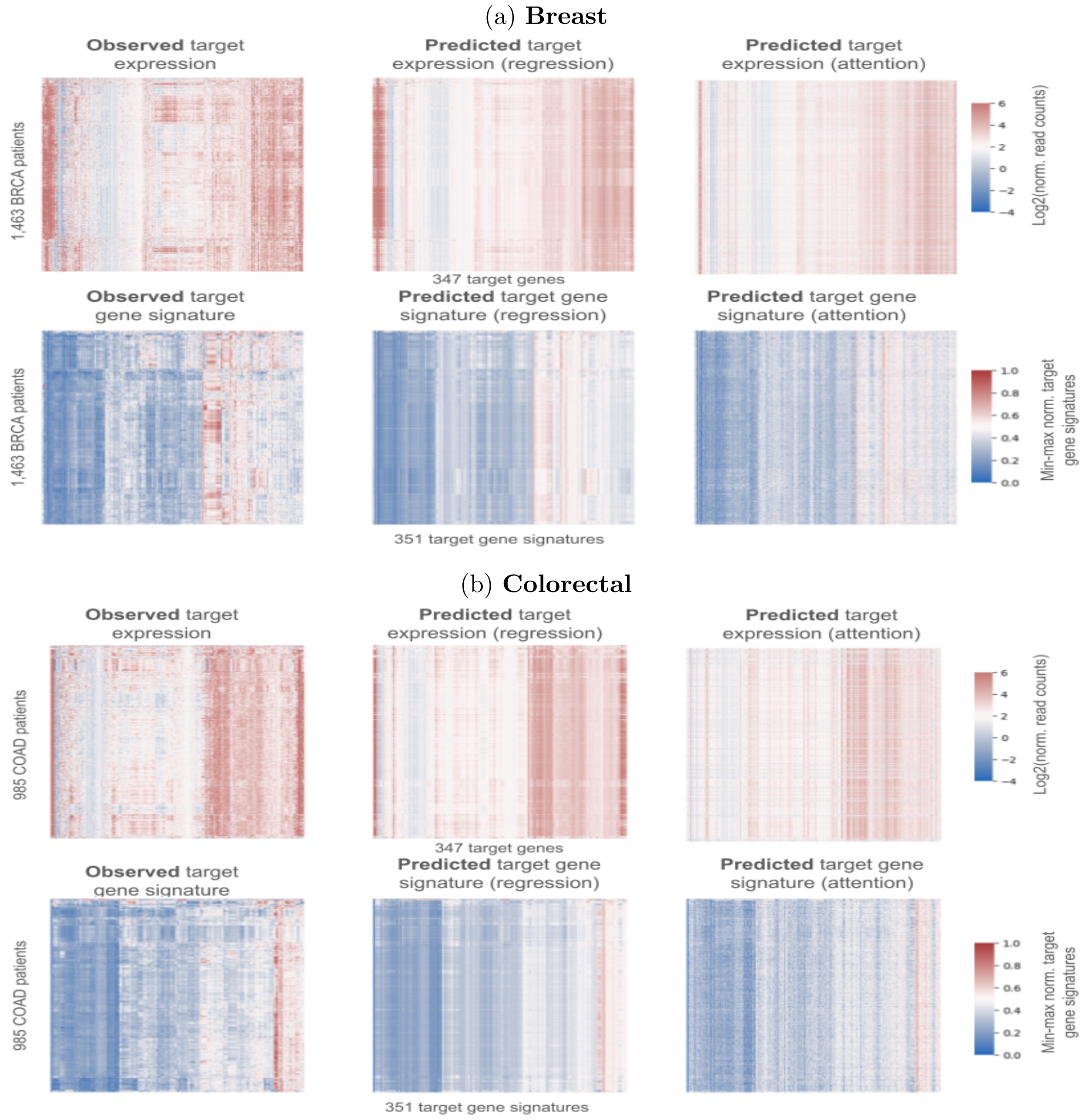

Figure 5: **Comparison of observed expression/signature matrices with those predicted on the basis of histopathology, stratified by cancer type.** Predictions are generated via cross-validation, such that a patient is not used to train the model that generates their predictions. Left is the observed gene expression or signature matrix. Center is the best-performing prediction model. Right is a spatially-aware model that includes tile-level transformer-based attention. The color bar on the left of each plot annotates cancer type.

##### 1.2.2 Binary classification

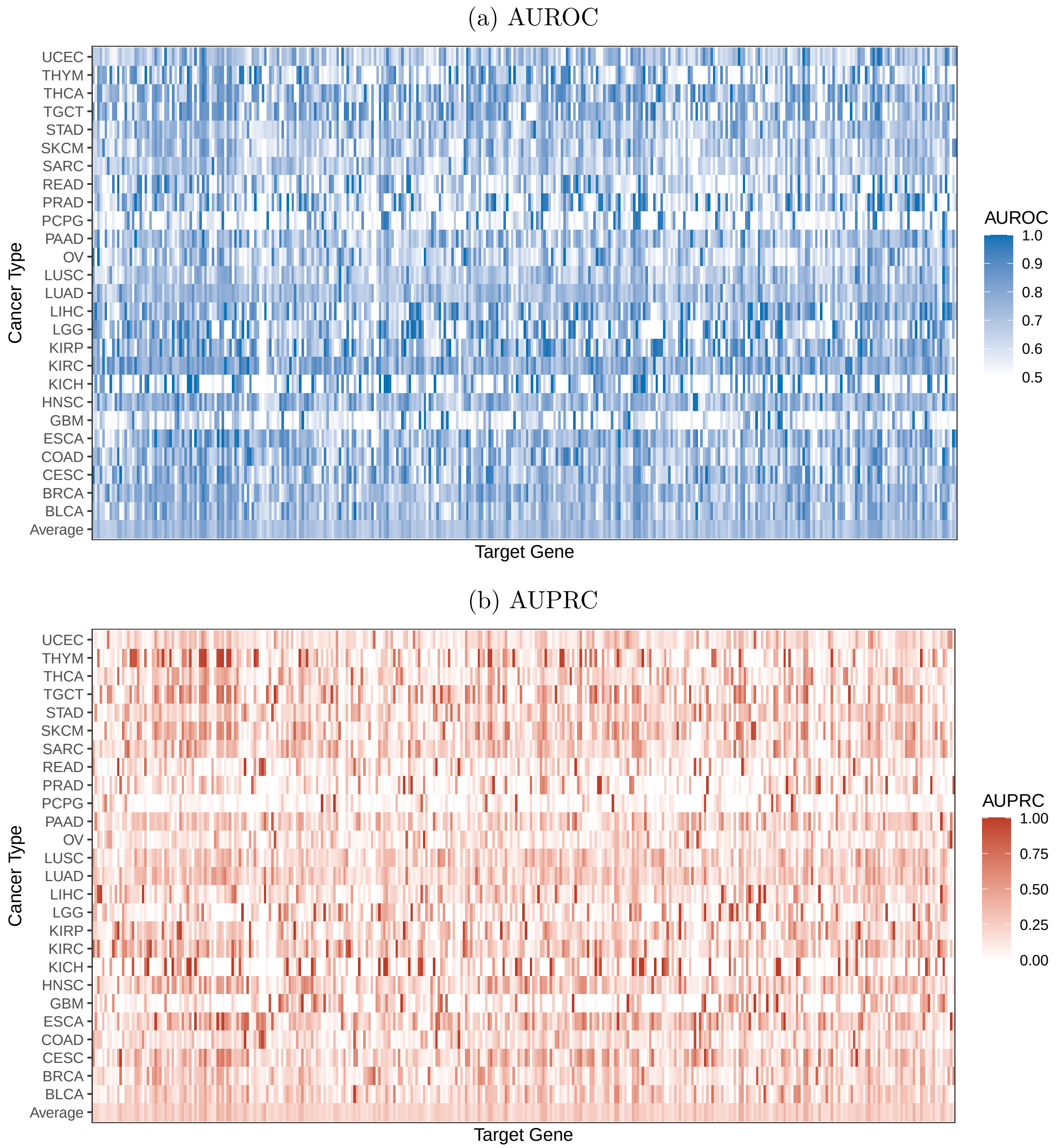

Figure 6: **AUROC and AUPRC of elevated target expression from digital histopathology, stratified by cancer type.** A patient is defined to have elevated expression if their expression level exceeds the 95th percentile for a given target. Performance is evaluated at the patient level in a held-out evaluation set and averaged across 8 cross-validation folds.

##### 1.2.3 Regression

(a) Pearson

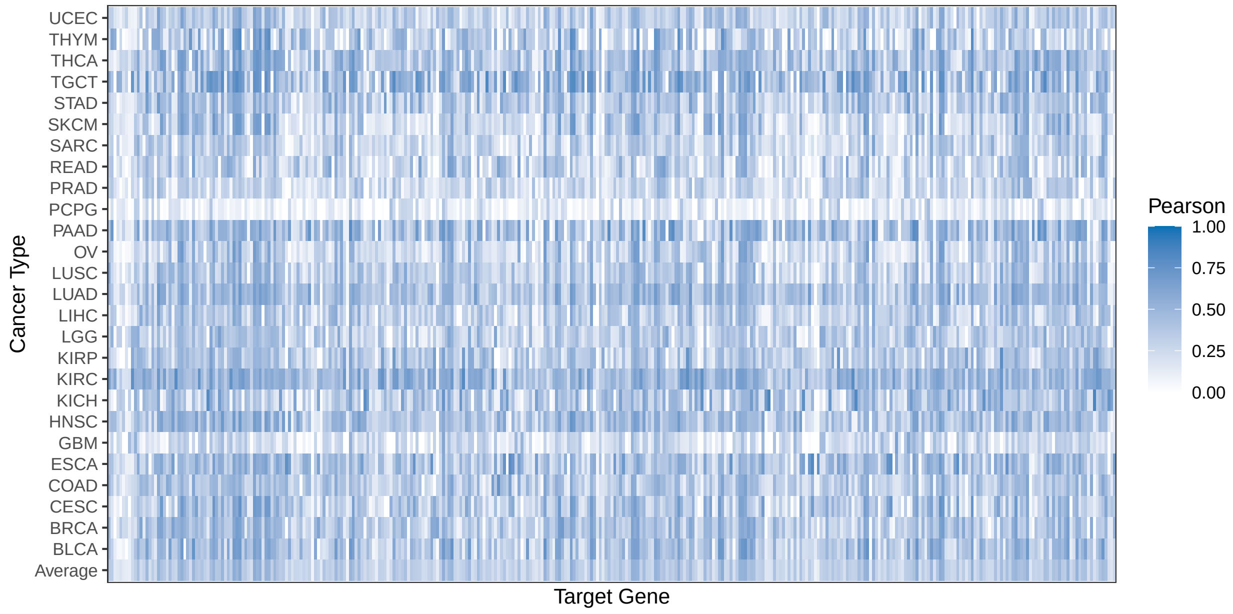

(b) Spearman

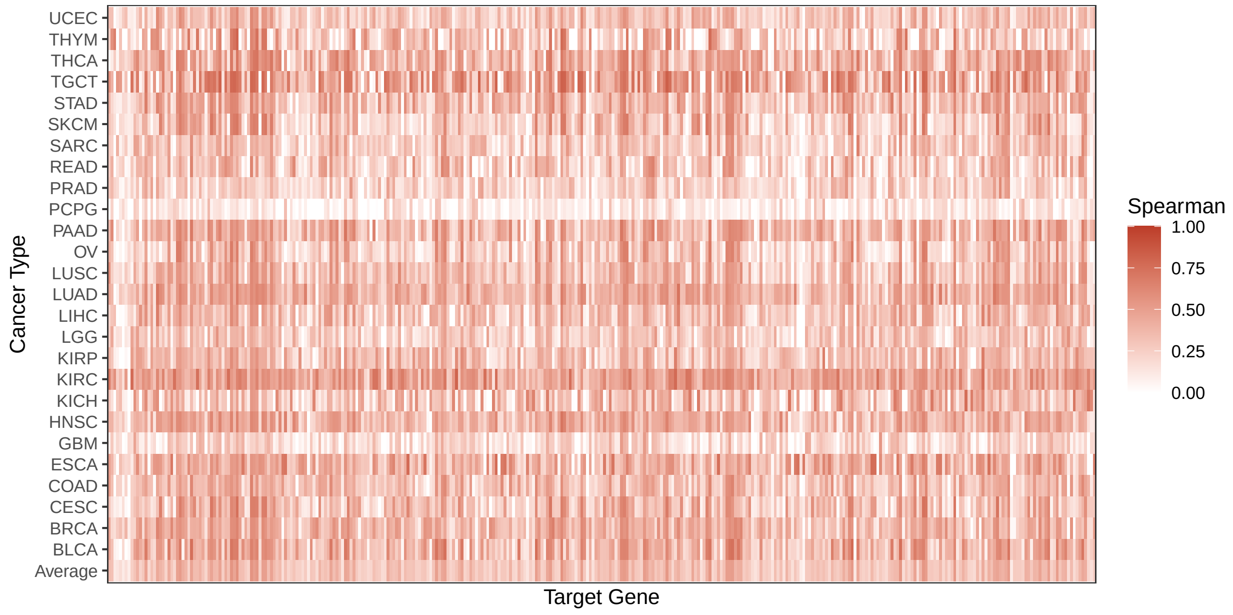

Figure 7: **Prediction of target expression level from digital histopathology, stratified by cancer type.** Performance is evaluated at the patient level in a held-out evaluation set and averaged across 8 cross-validation folds.

#### 1.3 Amplification Signature

##### 1.3.1 Binary classification

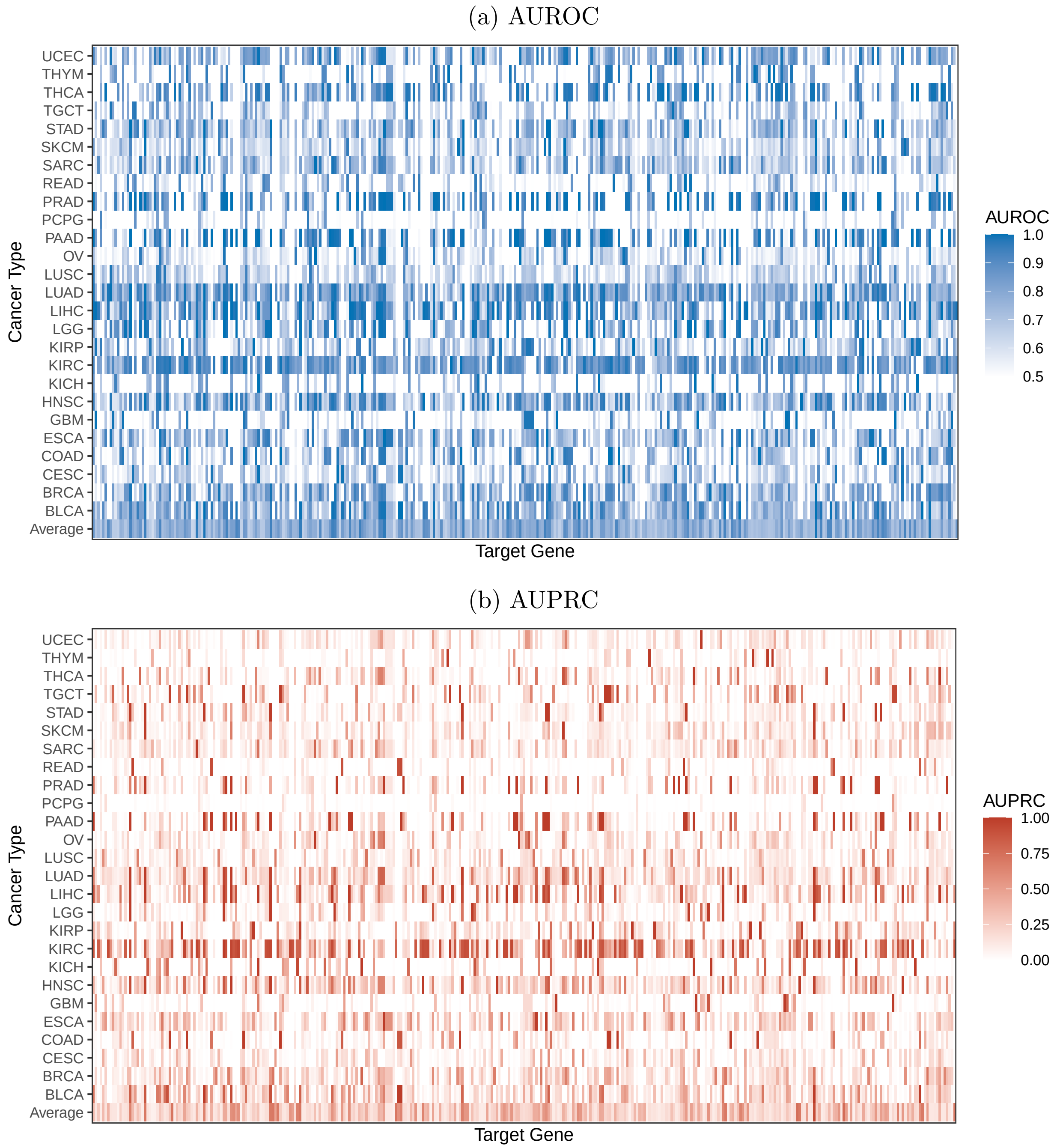

Figure 8: **Prediction of elevated amplification signature from digital histopathology, stratified by cancer type.** A patient is defined to have an elevated amplification signature if their score exceeded the 95th percentile for a given target. Performance is evaluated at the patient level in a held-out evaluation set and averaged across 8 cross-validation folds.

##### 1.3.2 Regression

(a) Pearson

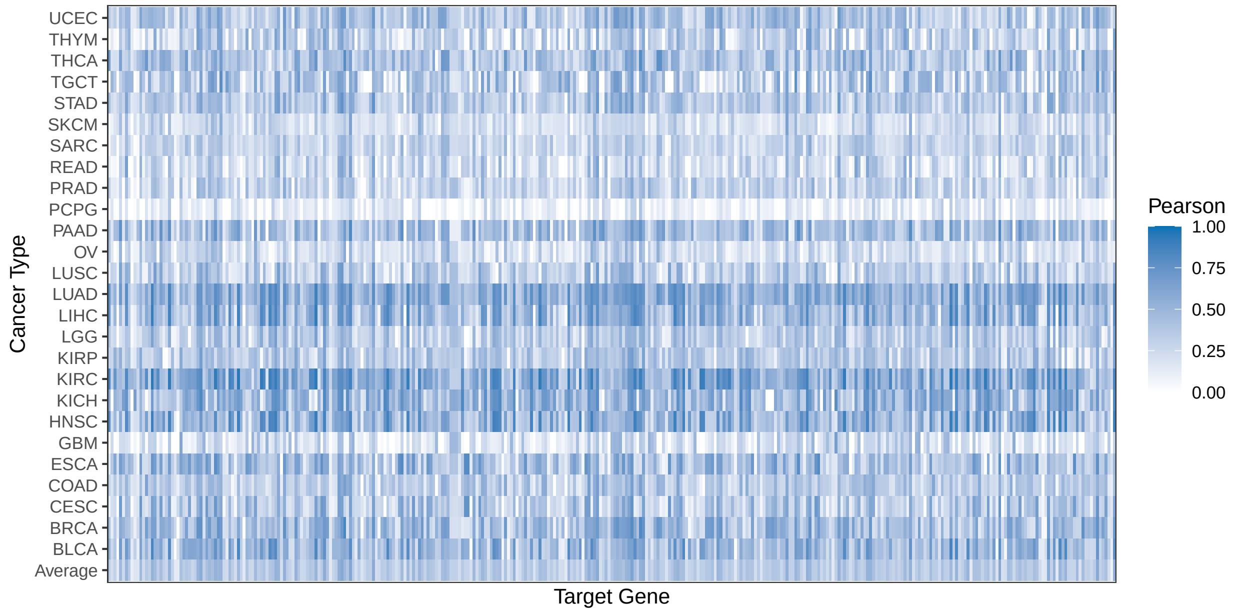

(b) Spearman

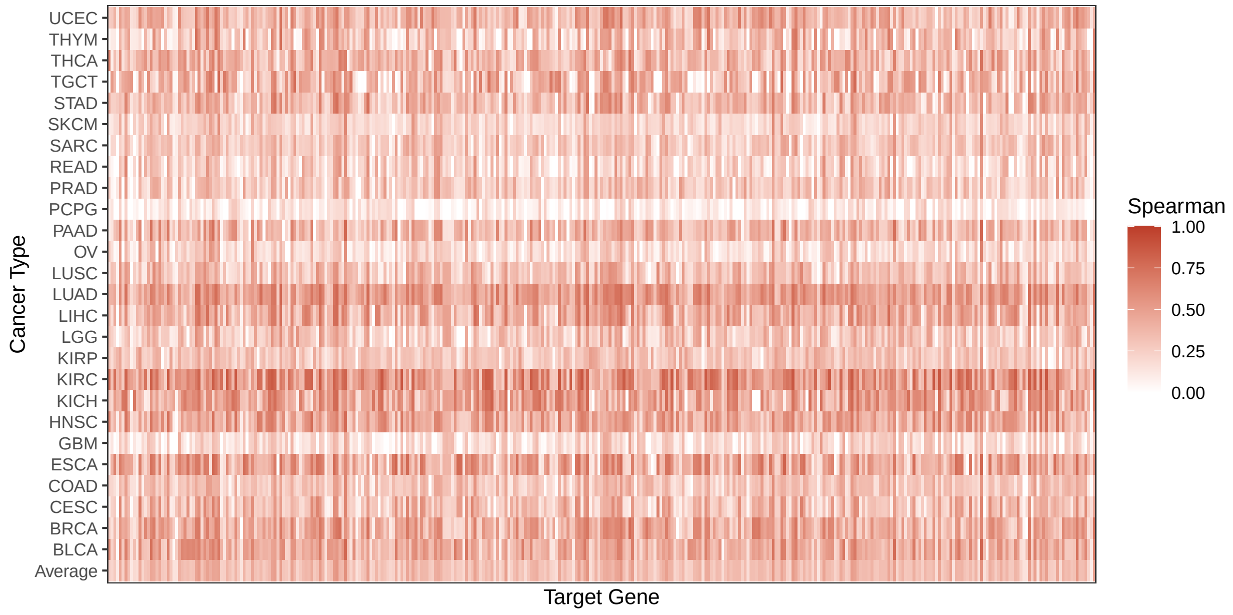

Figure 9: **Prediction of amplification signature from digital histopathology, stratified by cancer type.** Performance is evaluated at the patient level in a held-out evaluation set and averaged across 8 cross-validation folds.

#### 1.4 Cross Modality Comparisons

##### 1.4.1 Binary classification

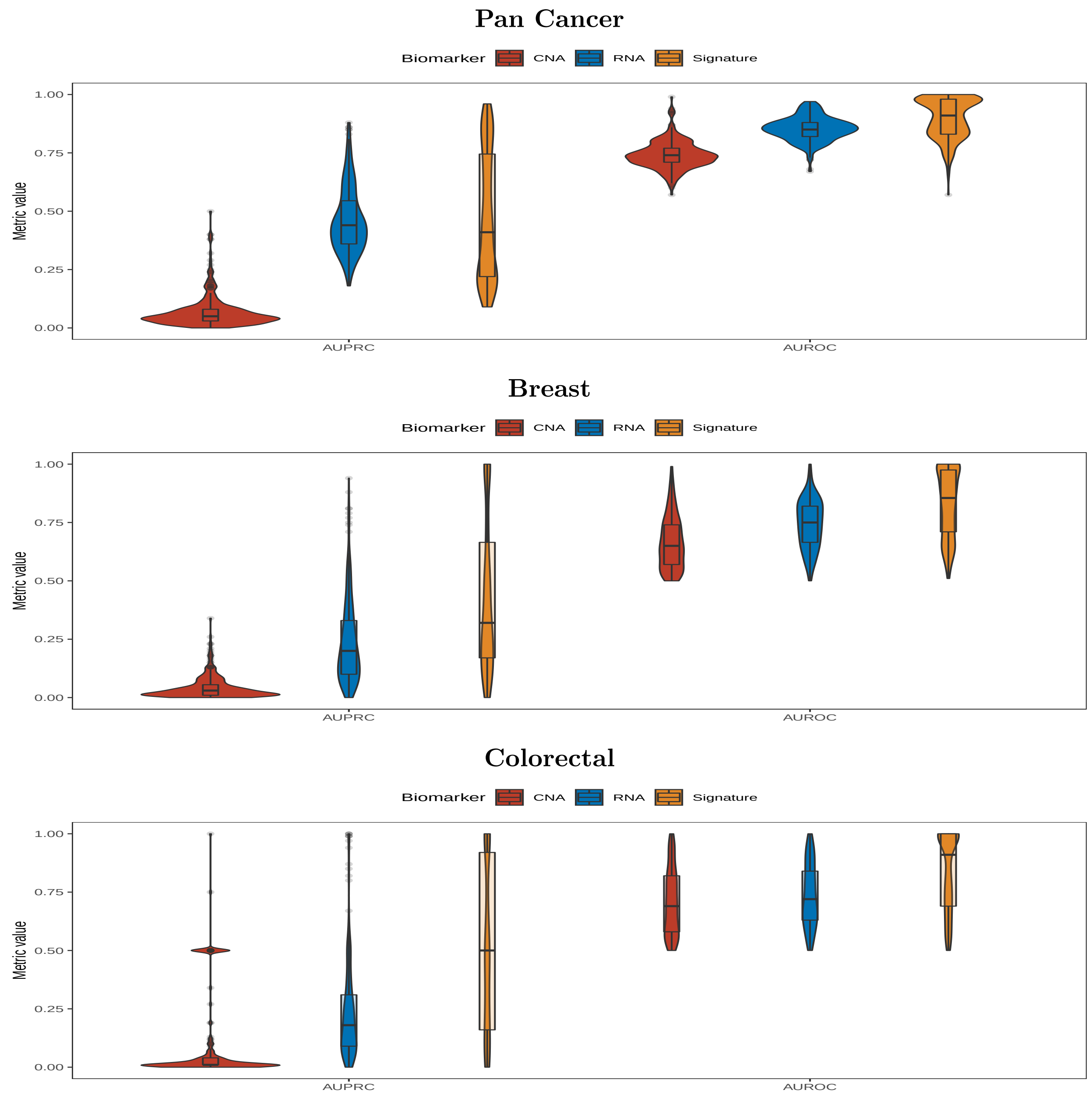

Figure 10: **Performance on the binary classification task, across biomarkers, stratified by cancer type.** The performance metrics are the areas under the precision-recall (AUPRC) and the receiver operating characteristic (AUROC). Performance is evaluated at the patient level in a held-out evaluation set. For copy number amplification (CNA), the task was to predict whether the patient harbored an amplification. For target expression (RNA) and the amplification signature, the task was to predict whether the patient's expression/signature level exceeded the 95th percentile. Distributions are shown across the 352 target genes.

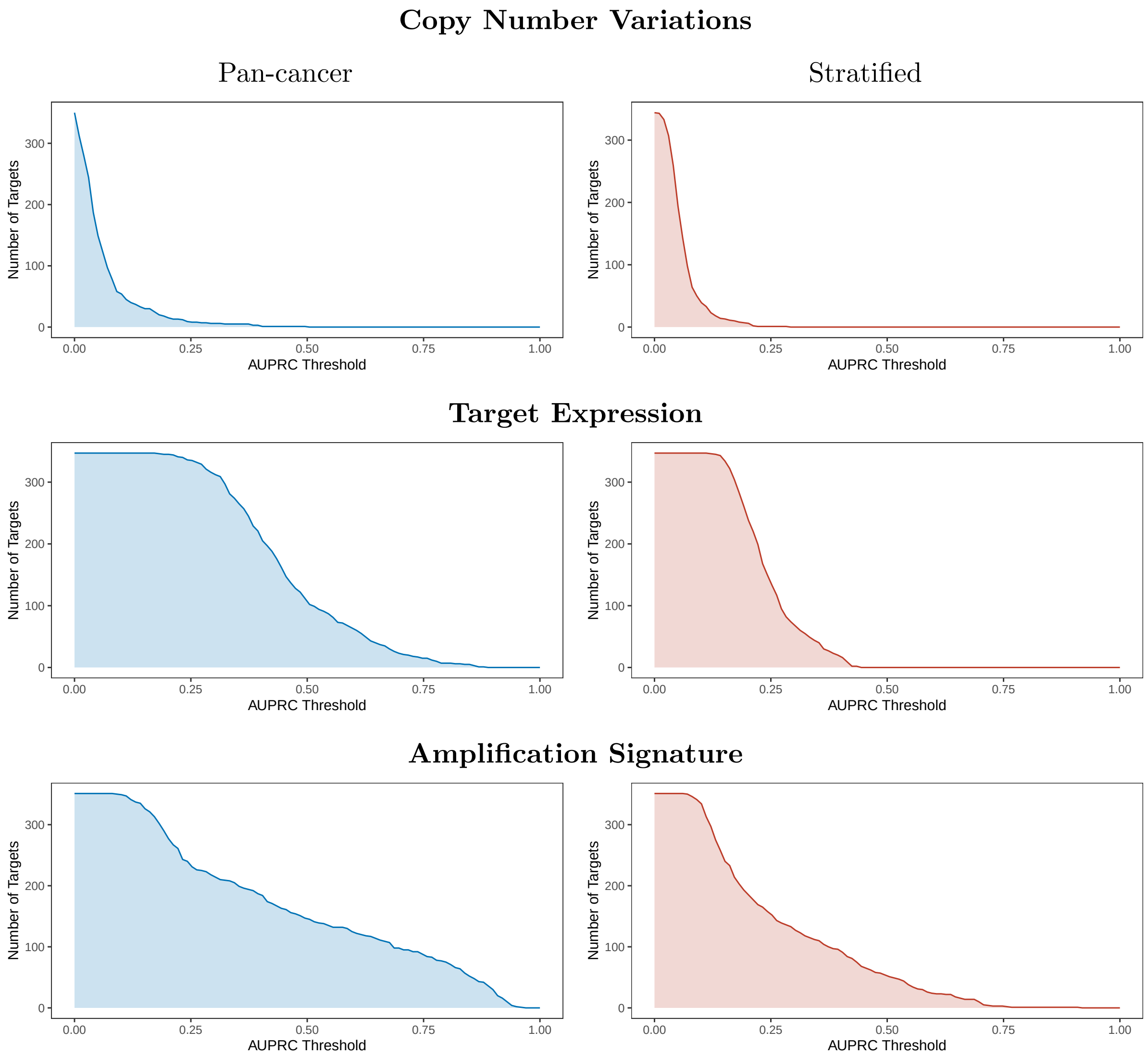

Figure 11: **Count of targets with AUPRC exceeding a given threshold for the pan-cancer and stratified binary classification task.** Performance is evaluated at the patient level in a held-out evaluation set and averaged across 8 cross-validation folds. For the pan-cancer evaluation, the AUPRC is calculated across all patients. For the stratified evaluation, the AUPRC is calculated separately within each cancer type, then averaged across cancer types. For copy number amplification, the task was to predict whether the patient harbored an amplification. For target expression and the amplification signature, the task was to predict whether the patient's expression/signature level exceeded the 95th percentile.

Table 1: **Count of genes with AUROC exceeding a given threshold.** Performance is evaluated at the patient level in a held-out evaluation set and averaged across 8 cross-validation folds. For the pan-cancer evaluation, the AUROC is calculated across all patients. For the stratified evaluation, the AUROC is calculated separately within each cancer type, then averaged across cancer types.

| Biomarker | Cohort | N | $N \geq 0.65$ | $N \geq 0.75$ | $N \geq 0.85$ | $N \geq 0.95$ |
|---|---|---|---|---|---|---|
| CNA | Pan-cancer | 352 | 334 | 142 | 11 | 0 |
| CNA | Stratified | 352 | 255 | 25 | 0 | 0 |
| RNA | Pan-cancer | 347 | 347 | 339 | 200 | 18 |
| RNA | Stratified | 347 | 311 | 97 | 0 | 0 |
| SIG | Pan-cancer | 351 | 349 | 335 | 246 | 145 |
| SIG | Stratified | 351 | 346 | 201 | 76 | 7 |

Table 2: **Count of genes with AUPRC exceeding a given threshold.** Performance is evaluated at the patient level in a held-out evaluation set and averaged across 8 cross-validation folds. For the pan-cancer evaluation, the AUROC is calculated across all patients. For the stratified evaluation, the AUROC is calculated separately within each cancer type, then averaged across cancer types.

| Biomarker | Cohort | N | $N \geq 0.1$ | $N \geq 0.2$ | $N \geq 0.3$ | $N \geq 0.4$ | $N \geq 0.5$ |
|---|---|---|---|---|---|---|---|
| CNA | Pan-cancer | 352 | 58 | 18 | 6 | 3 | 1 |
| CNA | Stratified | 352 | 39 | 7 | 0 | 0 | 0 |
| RNA | Pan-cancer | 347 | 347 | 345 | 316 | 221 | 112 |
| RNA | Stratified | 347 | 347 | 239 | 71 | 16 | 0 |
| SIG | Pan-cancer | 351 | 350 | 290 | 218 | 187 | 147 |
| SIG | Stratified | 351 | 335 | 185 | 131 | 92 | 52 |

##### 1.4.2 Regression

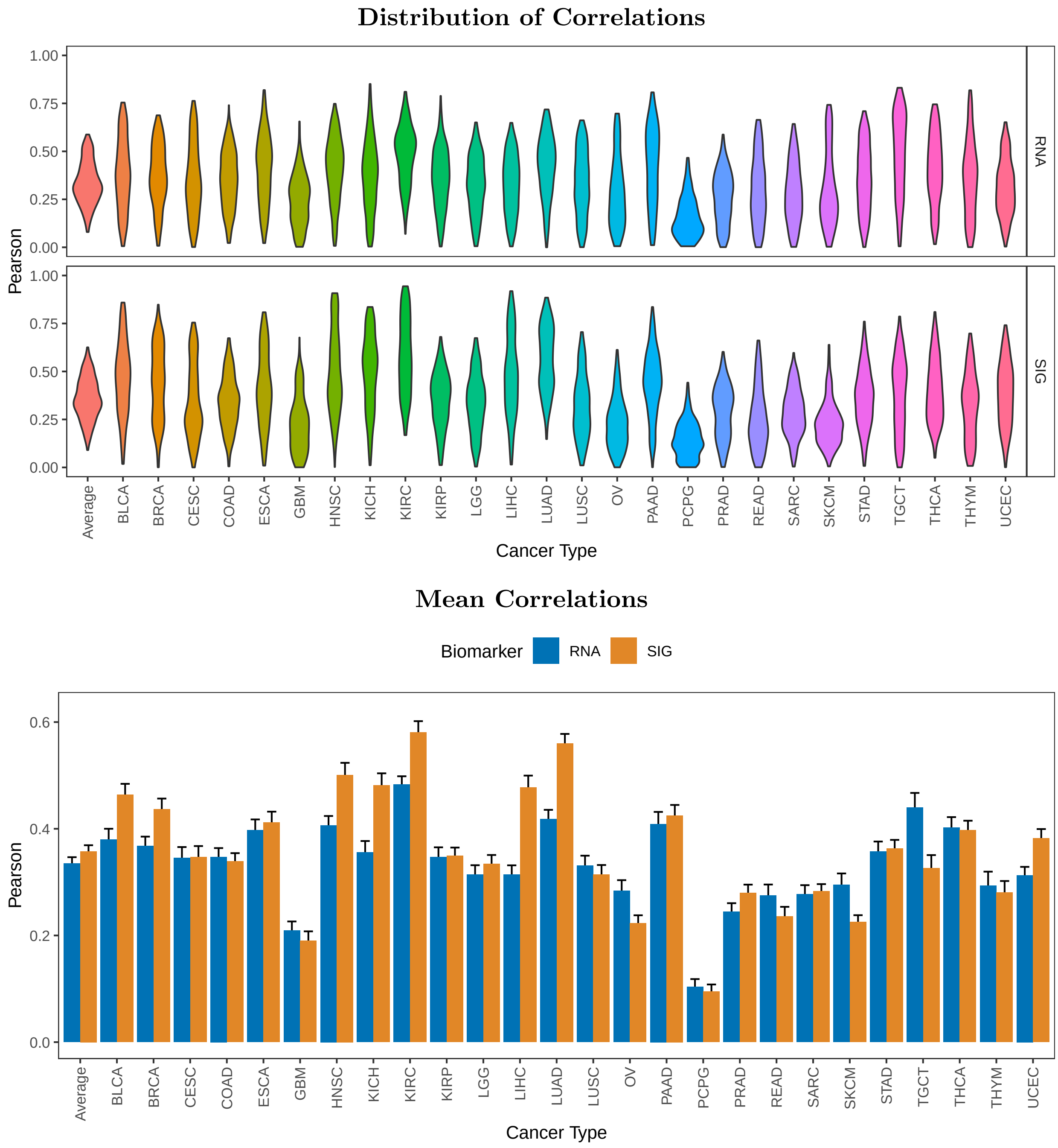

Figure 12: **Cross-modality comparison of continuous digital biomarker prediction quality, stratified by cancer-type.** Performance is evaluated at the patient level in a held-out evaluation set and averaged across 8 cross-validation folds. Metrics are calculated separately in each cancer type with $\geq 100$ patients. The distributions are shown across up to 352 target genes. For RNA, the task is to predict the normalized $\log_2$ expression level. For SIG, the task is to predict the min-max normalized amplification signature.

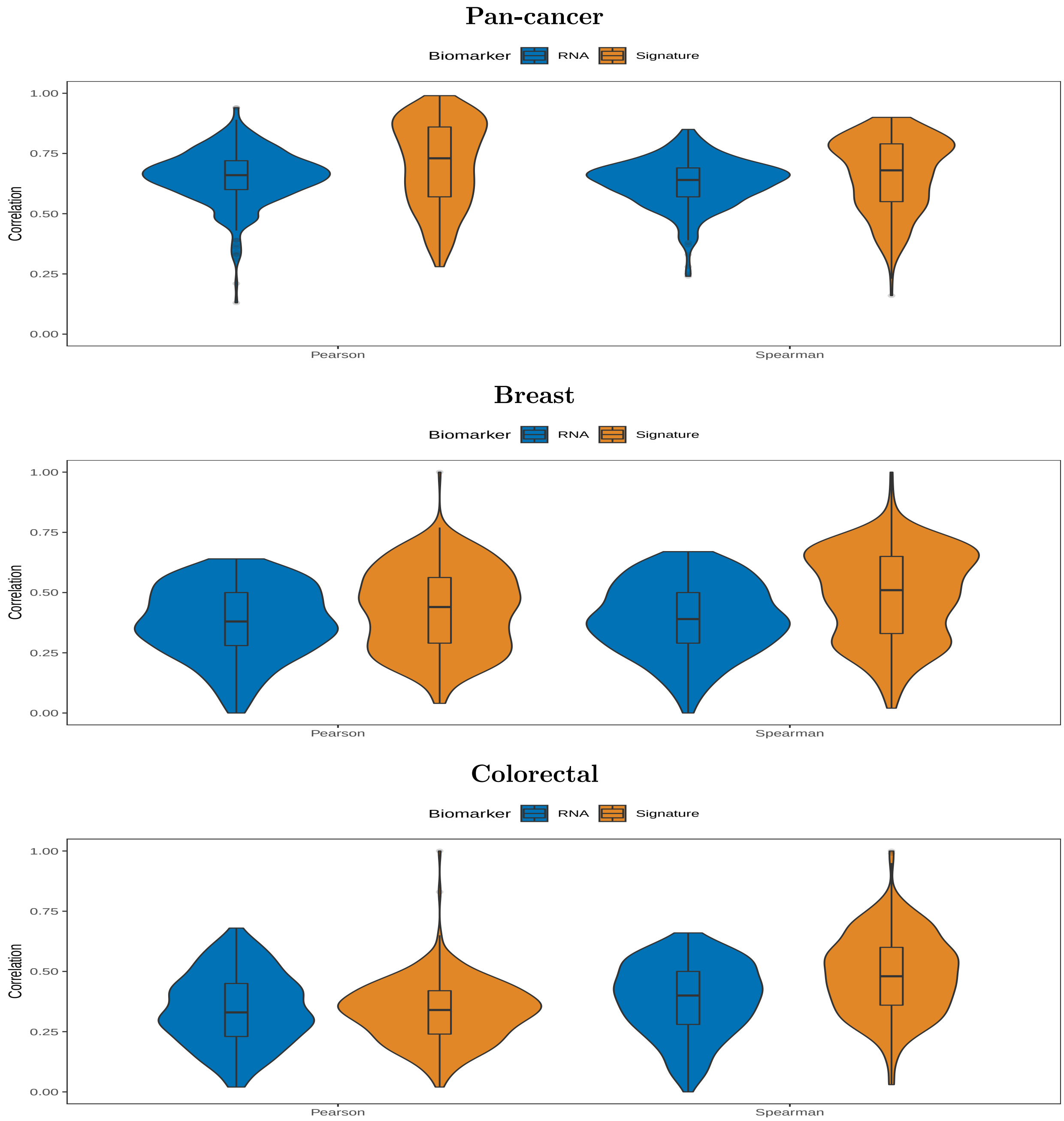

Figure 13: **Performance on the regression task, across biomarkers, pan-cancer and for two specific cancer types.** The task was to predict the continuous expression level or amplification signature score. Performance is evaluated at the patient level in a held-out evaluation set and averaged across 8 cross-validation folds. Distributions are shown across up to 352 target genes.

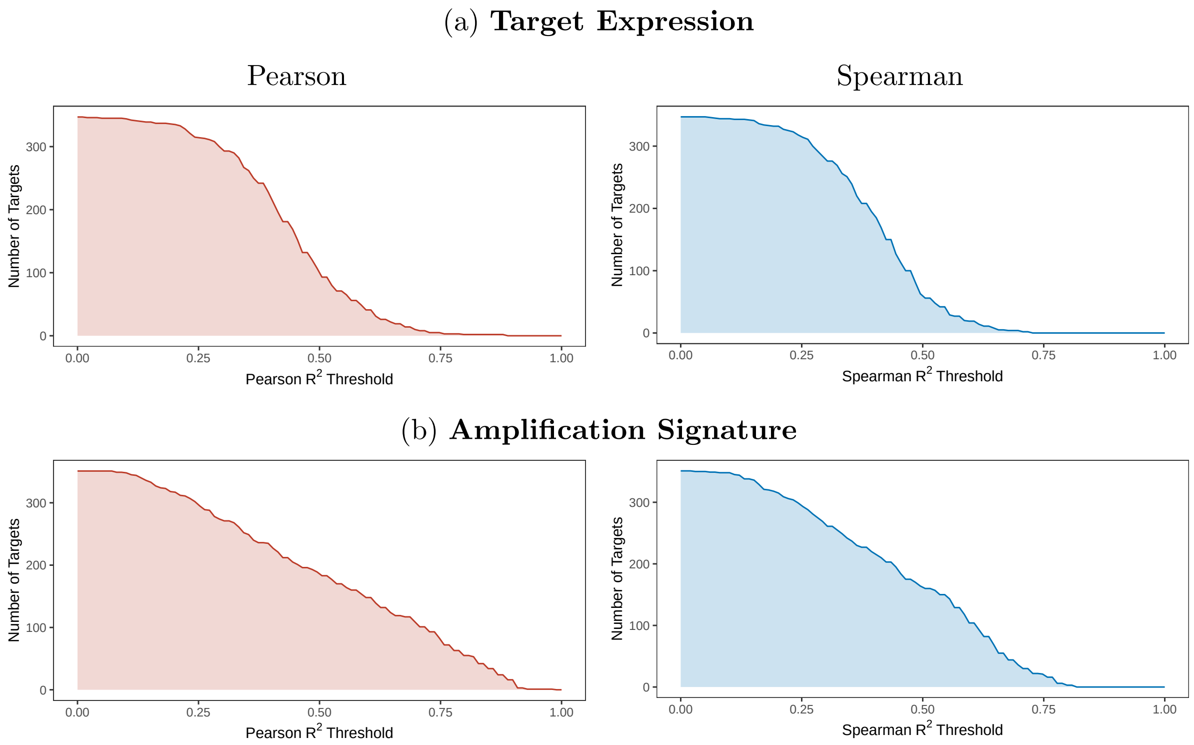

Figure 14: **Count of targets with Pearson and Spearman $R^2$ exceeding a given threshold for the pan-cancer regression task.** Performance is evaluated at the patient level in a held-out evaluation set and averaged across 8 cross-validation folds. Pearson and Spearman are calculated pan-cancer

(a) **Target Expression**

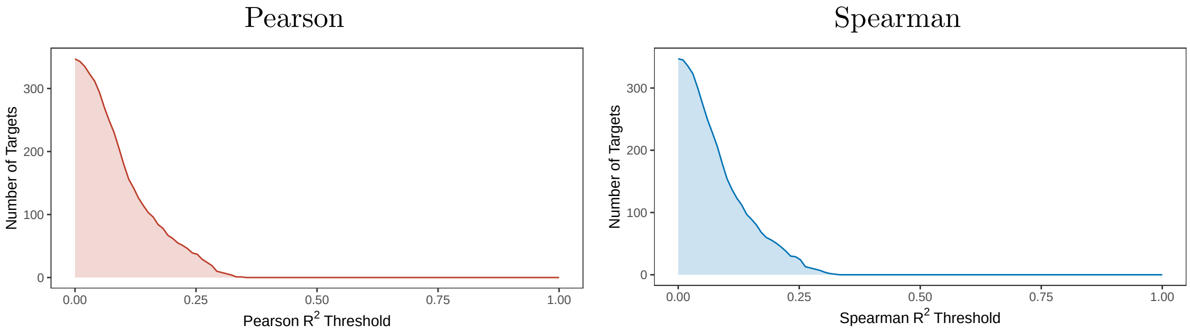

(b) **Amplification Signature**

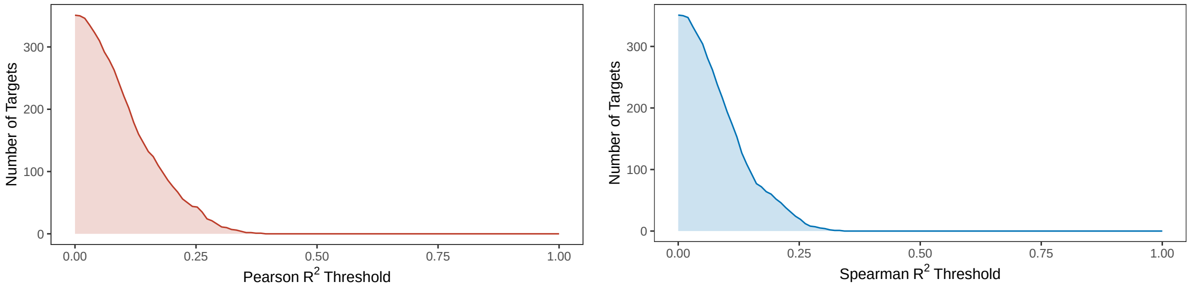

Figure 15: **Count of targets with Pearson and Spearman $R^2$ exceeding a given threshold for the stratified regression task.** Performance is evaluated at the patient level in a held-out evaluation set and averaged across 8 cross-validation folds. Pearson and Spearman are calculated separately in each cancer type with $\geq 100$ patients, then averaged across cancer types.

##### 1.4.3 Stratified

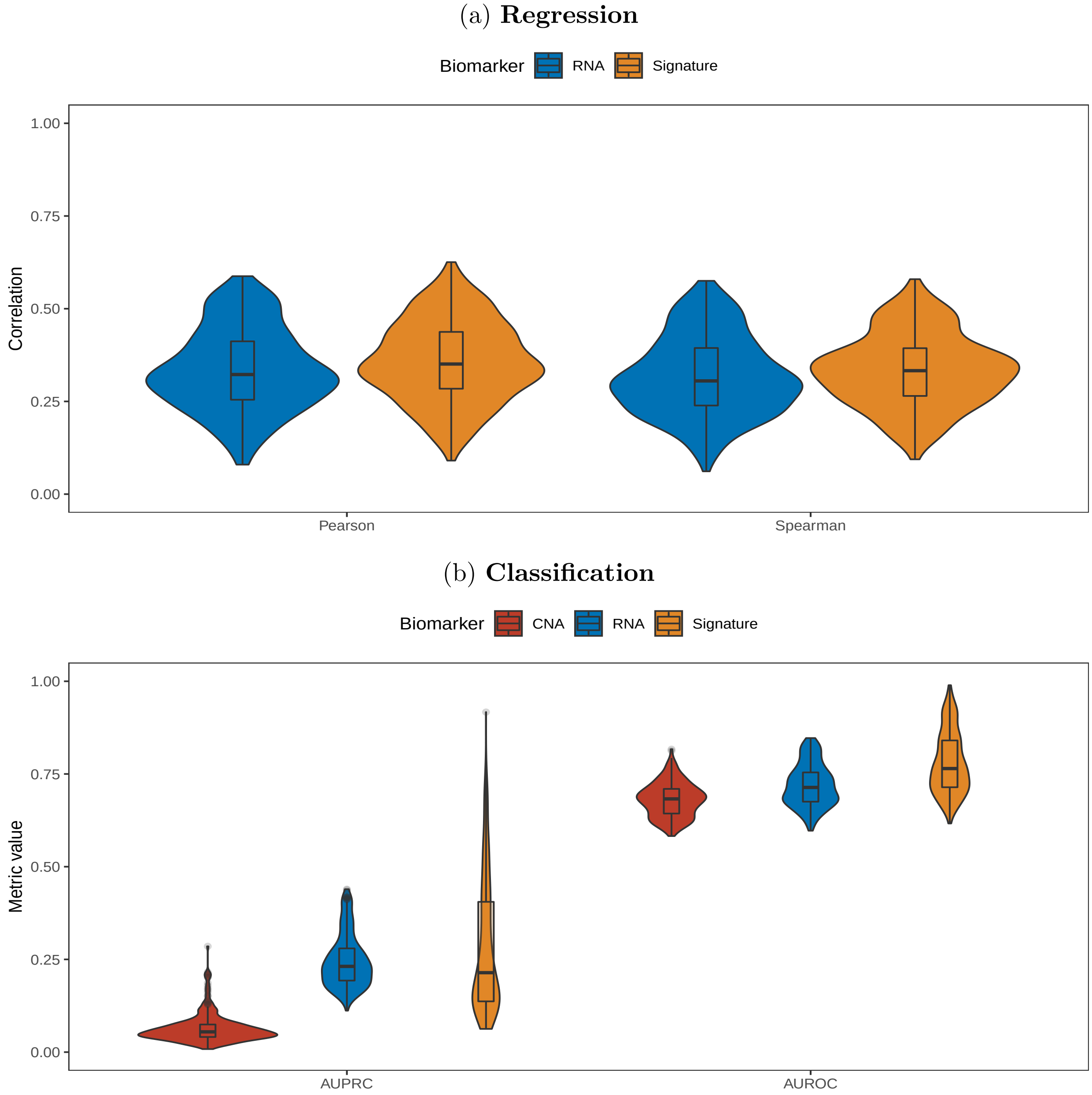

Figure 16: **Cross-modality comparison of digital biomarker prediction quality, stratified by cancer-type.** Performance is evaluated at the patient level in a held-out evaluation set and averaged across 8 cross-validation folds. Metrics are calculated separately in each cancer type with $\geq 100$ patients, then averaged across cancer types. The distributions are shown across up to 352 target genes. (a) Prediction of continuous target expression (RNA) and amplification signature levels. (b) Prediction of binary copy number amplification (CNA) status or being in the upper 5th percentile (RNA & signature). RNA expression is measured as $\log_2$ transcripts per million. Amplification signatures are based on those genes differentially expressed in patients with and without amplification.

#### 1.5 Amplification Signature Development

##### 1.5.1 Prevalence

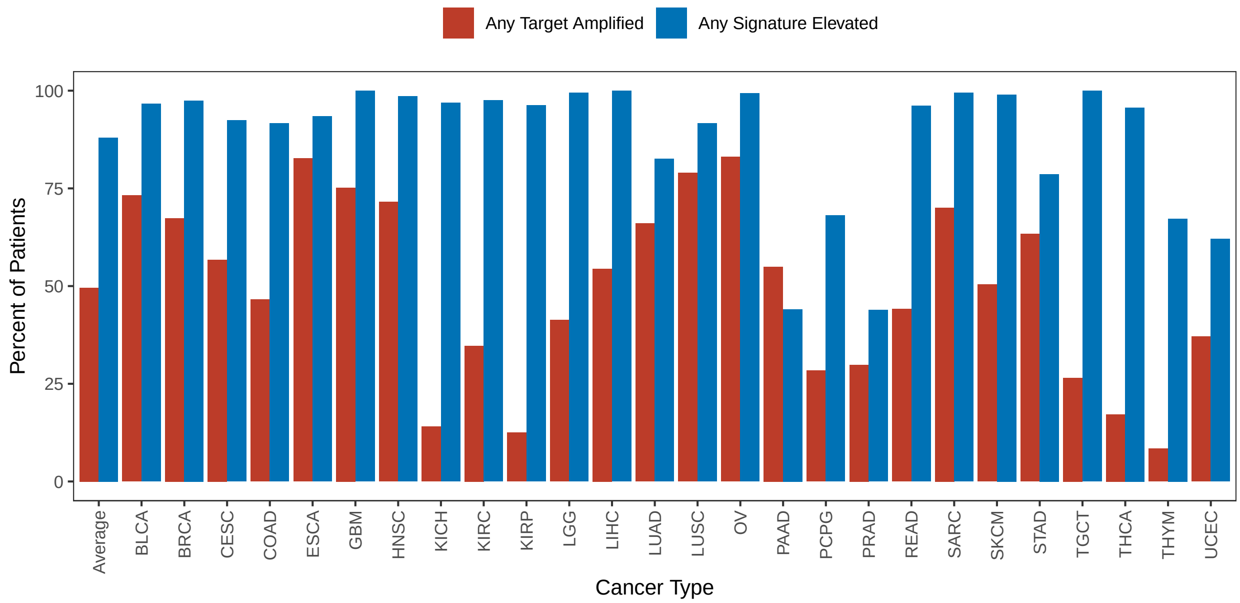

Figure 17: **Prevalence of any target amplified versus any amplification signature elevated stratified by cancer type.** Prevalence is calculated at the patient-level across up to 352 target genes. A patient was considered to have an elevated amplification signature if their score exceeded the 95th percentile for a given target.

##### 1.5.2 Signature by amplification

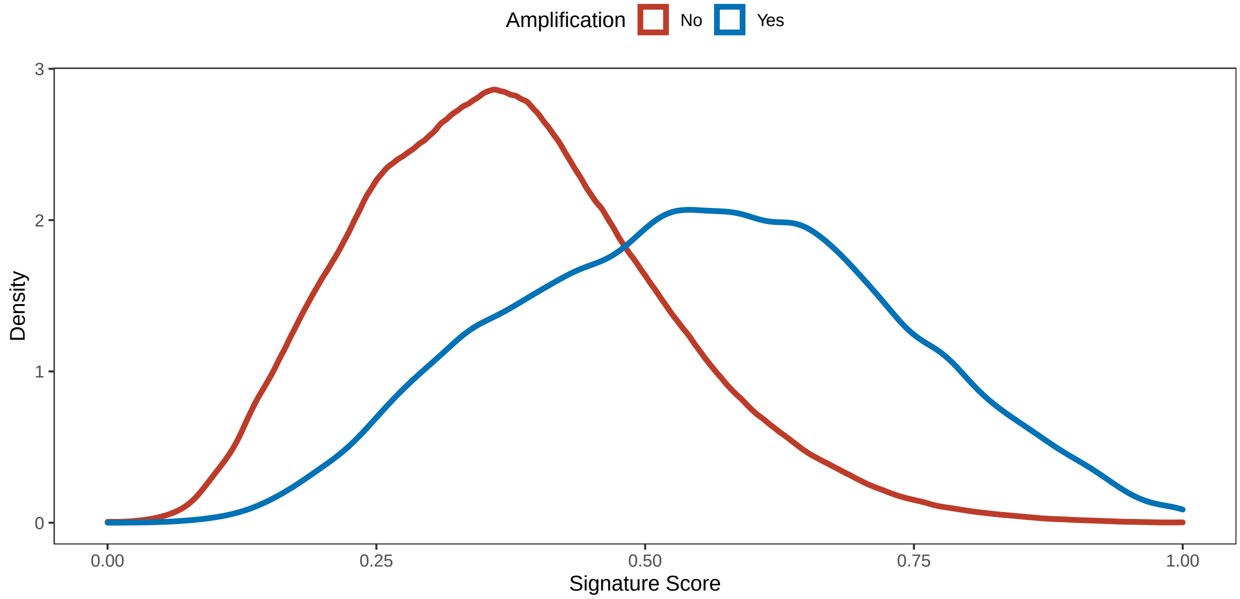

Figure 18: **Signature distribution by patient amplification status**. Shown is the average distribution across up to 351 amplification signatures.

| Patients | Mean Signature | Relative Change |
|---|---|---|
| Cases | 0.549 | +46.3% |
| Controls | 0.375 | |

Table 3: **Mean signature scores in patients with (cases) and without (controls) amplifications.** The mean is calculated across up to 351 amplification signatures.

##### 1.5.3 Correlation with expression

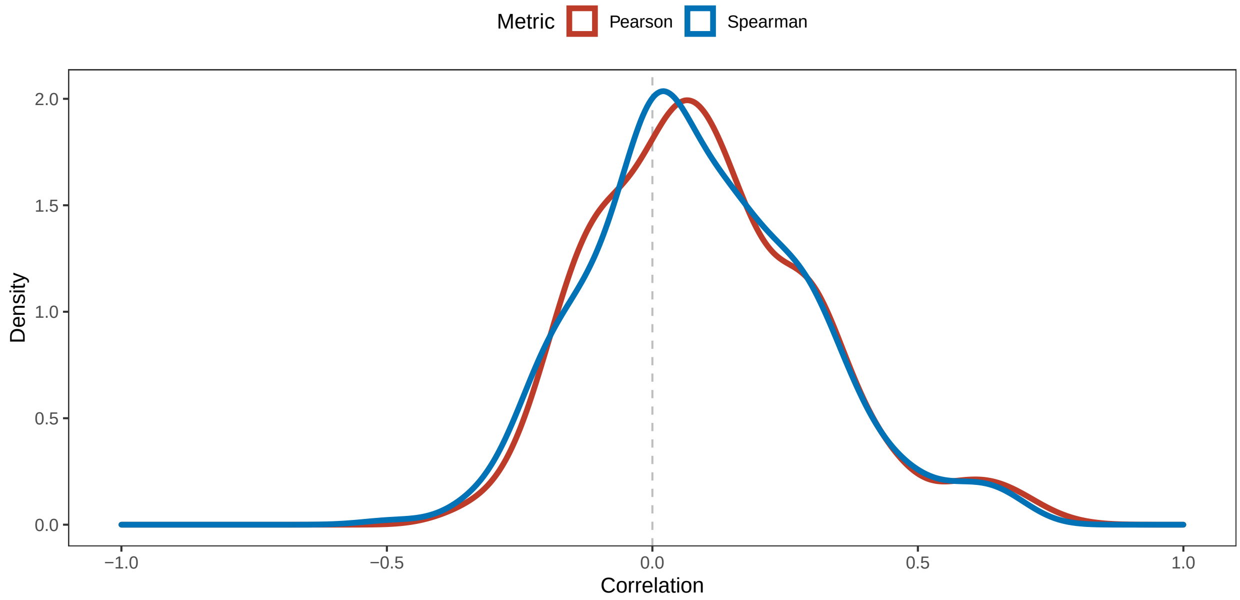

Figure 19: **Distribution of correlations between amplification signatures and expression of the amplified gene pan-cancer.** For each of 352 targets, the correlation is calculated across 14K subjects. Shown are the distributions of the 352 correlations.

| Metric | Min | Median | Mean | Max |
|---|---|---|---|---|
| Pearson $R^2$ | 0.000 | 0.018 | 0.053 | 0.568 |
| Spearman $R^2$ | 0.000 | 0.020 | 0.052 | 0.474 |

Table 4: **Squared correlation between the amplification signature and expression of the amplified gene, pan-cancer.** For each of 352 targets, the correlation is calculated pan-cancer. Shown are summary statistics for up to 352 correlations.

| Metric | Min | Median | Mean | Max |
|---|---|---|---|---|
| Pearson $R^2$ | 0.000 | 0.044 | 0.094 | 0.702 |
| Spearman $R^2$ | 0.000 | 0.045 | 0.090 | 0.671 |

Table 5: **Squared correlation between the amplification signature and expression of the amplified gene, stratified.** Correlations are first calculated within cancer types, then the mean is taken across cancer types. Shown are summary statistics for up to 352 correlations.

#### 1.6 Interpretability

##### 1.6.1 Breast cancer

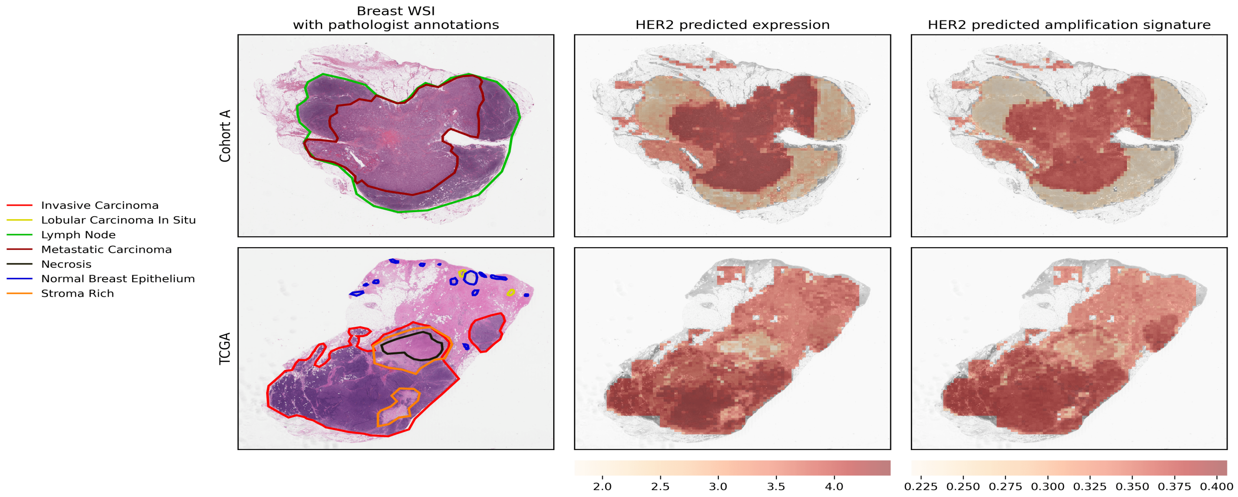

Figure 20: **Comparison of expression and signature predictions with expert pathologist annotations in breast cancer.** The pathologist was blinded to the predictions. Although the expression/signature models provide tile-level predictions, they were trained only on bulk, not spatially resolved, information.

##### 1.6.2 Colorectal cancer

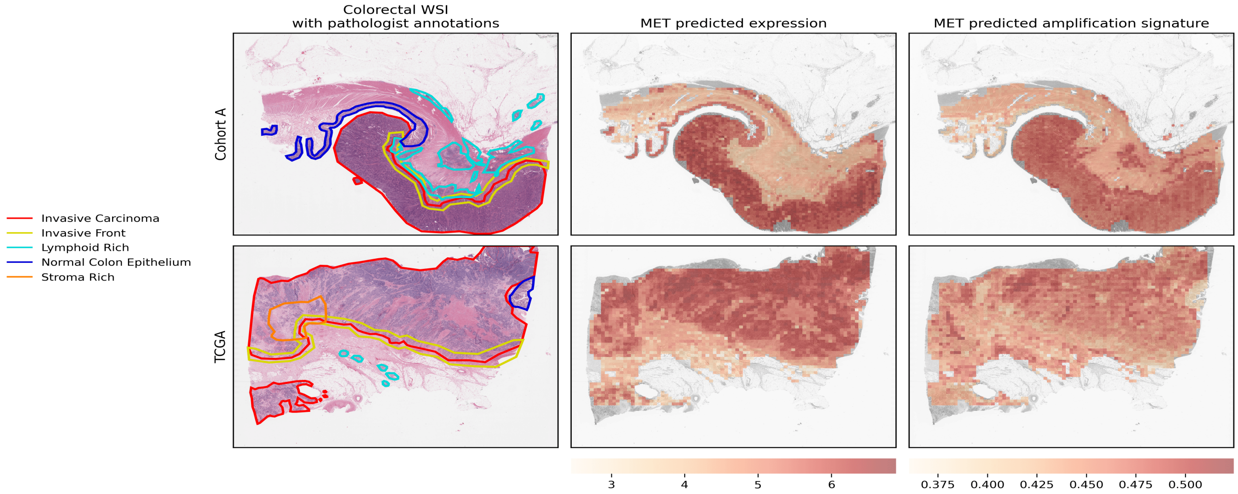

Figure 21: **Comparison of expression and signature predictions with expert pathologist annotations in colorectal cancer.** The pathologist was blinded to the predictions. Although the expression/signature models provide tile-level predictions, they were trained only on bulk, not spatially resolved, information.

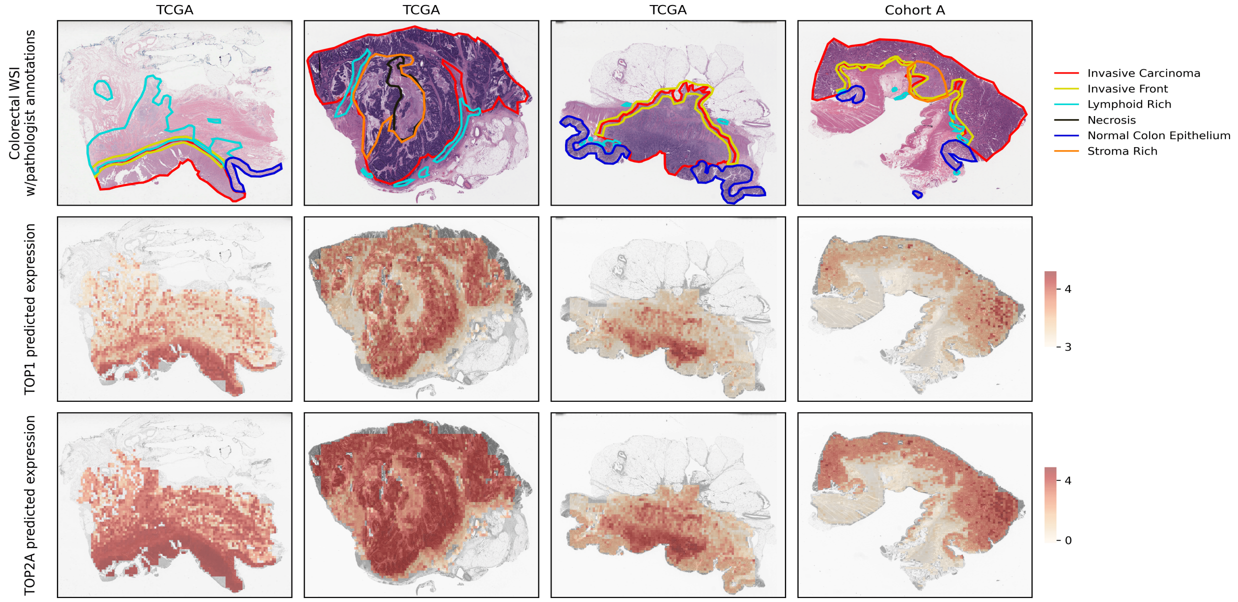

Figure 22: **Coexpression of TOP1 and TOP2A in colorectal cancer alongside pathologist annotations.** The pathologist was blinded to the predictions. Although the expression/signature models provide tile-level predictions, they were trained only on bulk, not spatially resolved, information.
